# Salivary DNA loads for human herpes viruses 6 and 7 are correlated with disease phenotype in Myalgic Encephalomyelitis/ Chronic Fatigue Syndrome

**DOI:** 10.1101/2021.01.06.20248486

**Authors:** Ji-Sook Lee, Eliana M. Lacerda, Luis Nacul, Caroline C. Kingdon, Jasmin Norris, Shennae O’Boyle, Chrissy H. Roberts, Luigi Palla, Eleanor M. Riley, Jacqueline M. Cliff

## Abstract

Myalgic Encephalomyelitis/ Chronic Fatigue Syndrome (ME/CFS) is a complex chronic condition affecting multiple body systems, with unknown cause, unclear pathogenesis mechanisms, and fluctuating symptoms which may lead to severe debilitation. It is frequently reported to have been triggered by an infection, particularly with herpes virus family members; however, there are no clear differences in exposure to, or seroprevalence of, any herpes virus in people with ME/CFS and healthy individuals. Herpes viruses exist in lytic and latent forms, and it is possible that ME/CFS is associated with viral reactivation, which has not been detectable previously due to insensitive testing methods.

Saliva samples were collected from 30 people living with ME/CFS at monthly intervals for six months and at times when they experienced symptom exacerbation, as well as from 14 healthy control individuals. The viral DNA load of the nine human herpes viruses was determined by digital droplet PCR. Symptoms were assessed by questionnaire at each time point.

Human herpes virus (HHV) 6B, HHV-7, herpes simplex virus 1 and Epstein Barr virus were detectable within the saliva samples, with higher HHV-6B and HHV-7 viral loads detected in people with ME/CFS than in healthy controls. Participants with ME/CFS could be broadly separated into two groups: one group displayed fluctuating patterns of herpes viruses detectable across the six months while the second group displayed more stable viral presentation. In the first group, there was positive correlation between HHV-6B and HHV-7 viral load and severity of symptom scores, including pain, neurocognition and autonomic dysfunction.

The results indicate that fluctuating viral load, related to herpesvirus reactivation state, may play a role in ME/CFS pathogenesis, or might be a consequence of dysregulated immune function. The sampling strategy and molecular tools developed permit large-scale epidemiological investigations.

**Contribution to the Field:** The cause of ME/CFS and the mechanisms underlying disease pathogenesis are not known, although symptoms are often triggered by infection. Human herpes virus (HHV) family members have been implicated, although there is no difference in the seroprevalence of any HHV in people with ME/CFS and healthy controls, showing there is similar prior infection rate. HHVs exist in either latent or active, lytic, phases in the human host, and it is possible that ME/CFS symptoms and their severity is related to HHV reactivation from a latent state. We have used droplet digital PCR, a sensitive and specific method, to measure the prevalence and DNA concentration of HHVs in the saliva of people with ME/CFS and controls, and analysed the correlation with disease over a six-month timecourse. We found that two HHVs, HHV-7 and HHV-6B, were elevated in saliva from people with ME/CFS, and that in people who were severely affected by ME/CFS, the concentration HHV DNA correlated with symptom severity over time in a subgroup of patients with fluctuating salivary HHV repertoire. Our study demonstrates the feasibility of measuring HHV concentration in readily acquired samples, enabling future large-scale studies aimed at testing the causal role of HHV reactivation in ME/CFS disease.

## 1. Introduction

Myalgic Encephalomyelitis/ Chronic Fatigue Syndrome (ME/CFS) is a disease of unknown aetiology, causing persistent or recurrent incapacitating fatigue; a hallmark symptom is post-exertional malaise (PEM). Other symptoms, which are present to a varying degree in different individuals, include pain and disturbances in immune function along with lymphadenopathy, unrefreshing sleep, cognitive difficulties, and dysfunction of the endocrine and autonomic nervous systems [1, 2]. Despite a prevalence in Europe ranging from 0.1% to 2.2% [3], there is still no diagnostic test for ME/CFS, hampering research into its cause and pathogenesis, and development of treatments. While the risk of developing ME/CFS is likely to be multifactorial, in approximately 50% of cases ME/CFS symptoms appear after a “viral-like” illness, and it is possible that an infectious disease can trigger ME/CFS onset. However, no single causative pathogen has been identified for ME/CFS, despite extensive research [4].

Although there have been reports that bacterial infections such as Lyme disease [5] or Q fever [6] can trigger ME/CFS, it is mostly commonly viral infections, especially those from the herpes virus family, which have been more widely associated with ME/CFS [4]. There are nine members of the human herpes virus (HHV) family which naturally infect humans: herpes simplex virus-1 and −2 (HSV-1, HSV-2), varicella-zoster virus (VZV), which is the causative agent of chicken pox and shingles, Epstein-Barr virus (EBV), which causes of infectious mononucleosis, human cytomegalovirus (HCMV), HHV6 including subtypes HHV6A and HHV6B, HHV7 and Kaposi’s sarcoma virus (KSHV). These viruses are all of potential interest in ME/CFS due to their ability to latently infect either neurons or immune system cells, both of which may be affected in ME/CFS. There has been speculation that Epstein Barr Virus (EBV) is involved in ME/CFS pathogenesis for many years [7, 8], although reports are inconsistent [9, 10]. Reports of the potential involvement of CMV in ME/CFS are also mixed, with higher anti-CMV antibodies reported in serum in ME/CFS compared to healthy controls (HCs) in one study [11] but no correlation in another [12]. HHV-6 and HHV-7, which are from the *Roseolovirus* genus of the *Beta-herpesvirinae* subfamily, cause widespread long-term persistent infection with reported population prevalence of >90% [13] with infection usually occurring within the first three years of life. In early studies, there was serological indication of HHV-6 reactivation with higher specific anti-HHV-6 serum antibody concentration in people living with ME/CFS than in HCs [14, 15] but these data have been contradicted in other studies [16]. Similarly, HHV-7 infection is commonly detectable in people living with ME/CFS, but the detection rate of HHV-7 DNA is similar in ME/CFS patients and in HCs [17].

Thus, reports of association between herpes virus infection and ME/CFS are inconsistent. This may be due partly to small sample sizes in some studies, and to heterogeneity within and between ME/CFS populations included in different studies. We did not find any difference in seroprevalence or antibody titres against the eight herpes viruses between people living with ME/CFS and HCs in our previous work [18]. However, a hallmark feature of herpes viruses is their life cycle in which, following an acute infection, they remain in the host in a latent persistent form, and can reactivate following immune system disturbance. Measurements of antibody titre by ELISA and qPCR analysis of viral DNA concentration in blood may be too insensitive to fully distinguish reactivation events from latent infection, and a more sensitive analysis might enable the full characterisation of herpes virus reactivation events in ME/CFS, leading to a definitive answer regarding the role of herpes virus infections in this condition. Digital droplet PCR (ddPCR) permits absolute quantification of nucleic acids [19]. This is achieved via an emulsion process in which the fluorescent probe-based PCR reaction is partitioned into approximately 15,000 highly uniform, one nanolitre volume, reverse (water-in-oil) micelles that are stable at high temperatures: each droplet is essentially an independent nano-PCR. Enumeration of the number of positive and negative droplets post-PCR gives a sensitive and precise readout of the template DNA concentration.

In this study, we hypothesised that ME/CFS may follow an acute infection with a herpes virus, with symptoms being triggered by a virus reactivation, followed by a state of “aberrant homeostasis” [20]. Although these viral infections are common, it is possible that people living with ME/CFS experience prolonged or more frequent reactivation events than healthy individuals and are unable to consistently contain a latent infection. We aimed to develop highly sensitive and specific ddPCR assays for the human herpes family viruses, and use them to test for correlation between viral load in saliva, an accessible sample likely to be involved in transmission [16], and the clinical manifestations of disease over time, particularly during disease exacerbation.

## 2. Materials and Methods

### 2.1 Study Participants

Potential ME/CFS study participants were recruited to the UK ME/CFS Biobank (UKMEB) [21] through the UK National Health Service or referred via patient support groups with ME/CFS. They were required to have a confirmed medical diagnosis of ME/CFS and were re-assessed by clinical research staff to ensure compliance with the Canadian Consensus [22] and/or CDC-1994 (“Fukuda”) [1] criteria, which were the study case definitions. Non-fatigued HC participants were recruited as friends or family of people with ME/CFS or by advertisement in Higher Education Institutions and GP practices. Participants were aged between 18 and 60 years and provided written informed consent. Exclusion criteria included having taken anti-viral or immunomodulatory drugs within the previous three months, chronic comorbid disease, a history of infectious disease such as tuberculosis or hepatitis B or C, pregnancy and morbid obesity. Inclusion criteria included HHV DNA detected in either plasma or PBMC (DNA positive) or HHV DNA not detected in either sample (DNA negative (as described below, 5.2). Ethical approval was granted by the London School of Hygiene & Tropical Medicine (LSHTM) Ethics Committee (Ref. 6123) and the National Research Ethics Service (NRES) London-Bloomsbury Research Ethics Committee (REC ref. 11/10/1760, IRAS ID:77765). Peripheral Blood Mononuclear Cells (PBMC) and plasma samples were cryopreserved and stored from all UKMEB participants [21] and accessed for this study.

### 2.2 Longitudinal Analysis

Selected participants from the UKMEB cohort provided further written informed consent for this longitudinal study. Participants were sent GeneFiX™ Saliva 2ml DNA Collection tubes containing DNA stabilisation buffer and asked to return filled saliva tubes by post monthly for six months as well as on the first, third and fifth day of any disease exacerbation episode including acute illness or significant worsening of ME/CFS symptoms.

Clinical symptom assessment forms (Supplementary Figure 1), containing 58 questions with graded answers, were completed and returned at the same time as each saliva sample. From these data, symptom subgroup scores were developed, for immune, neuroendocrine, autonomic, neurocognition, pain, sleep, and post-exertional related symptoms; the Canadian Consensus Criteria assesses these domains for ME/CFS diagnosis [22]. The scores ranging from “0” to “100” represent a weighted average of the reported symptoms on each domain, which are reported as “absent, mild, moderate, or severe”.

### 2.3 DNA extraction and Digital Droplet PCR

DNA was extracted from stored plasma samples using the QIAamp MinElute Virus Spin Kit (Qiagen), and from PBMC and saliva using the QIAamp DNA Mini Kit (Qiagen), according to the manufacturer’s instructions. Each DNA extraction run included a seronegative control sample and a Clinical Virology Multiplex I (National Institute for Biological Standards and Control (NIBSC)) positive extraction control: this comprised Adenovirus serotype 2, BK virus, CMV, EBV, HSV-1, HSV-2, HHV-6A, HHV-6B, JC virus, Parvo B19 virus and VZV.

For assay development and to generate standard curves, titrated commercial quantitated DNA was used: HSV-1, HSV-2, VZV & HHV-8 were purchased from Vircell (Granada, Spain), HHV-6A, HHV-6B and HHV-7 were from ABI (Advanced Biotechnologies Inc, MD, USA) while EBV and CMV were from the National Institute for Biological Standards and Control (NIBSC).

Each 20µl ddPCR reaction mix contained 10 µl ddPCR™ Supermix for Probes (Bio-Rad Laboratories, Hercules, USA), 9 µl of template DNA, 1 µl of 20X primer and probe mix at a final concentration 900 nM (for primer) and at 250 nM (for probe). Positive (commercial viral DNA) and negative (water) controls were included in every ddPCR run. Primer and probe sequences are shown in Supplementary Table 1, and were obtained from publications [23-28]. The reaction mix was partitioned in oil-in-water with 70 µl of droplet generator oil using a QX-100 droplet generator (Bio-Rad). The generated droplets (around 40 µl) were transferred to ddPCR™ 96-Well Plates (Bio-Rad) using a multichannel pipette and covered with a pierceable PCR plate seal using a PX1™ PCR Plate Sealer (Bio-rad). PCR amplification was performed with the following thermal cycling: 95 °C for 10 min, 40 cycles consisting of 94 °C for 30 sec (denaturation) and 60 °C for 1 min (extension), followed by 98°C for 10 min and holding at 12 °C. After that, droplets (positive and negative) were analysed using a QX100 droplet reader (Bio-rad) and the target DNA concentration was calculated and presented as copies/µl in the reaction mixture using QuantaSoft software (Bio-Rad). The ddPCR result was determined as positive when the DNA samples showed at least three positive droplets from 10,000 to 15,000 droplets, according to the manufacturer’s recommendation.

### 2.4 Herpes virus serology

Commercial ELISA kits were used to measure plasma concentrations of IgG to HSV-1, HSV 2, VZV, HCMV, EBV viral capsid antigen (VCA), EBV nuclear antigen 1 (EBNA 1) (from Demeditec Diagnostics (Kiel, Germany)) and HHV-6 (from VIDIA (Vestec, Czech republic)): these data have been reported previously [18], and are included here for comparison. The antibody concentrations were calculated from standard curves. For qualitative evaluation of IgG antibodies to HSV-1, HSV-2, VZV, CMV, EBV VCA, EBV, EBNA-1, the concentration was interpreted as: concentration ≤ 8 U/ml = negative, concentration ≥ 12 U/ml = positive, and concentration between 8 and 12 U/ml = equivocal. For qualitative evaluation of IgG antibodies to HHV-6, concentration ≤10.5 U/ml was considered negative, while concentration ≥ 12.5 U/ml was considered positive and concentration between 10.5 and 12.5 was considered equivocal.

### 2.5 Data analysis

Statistical analysis was conducted using GraphPad Prism 8.4.2 (GraphPad Software, San Diego, CA). Continuous variables (viral loads, viral persistency and symptom scores) were described using median and interquartile ranges (IQR) values or min to max as indicated. Mann-Whitney test was used for viral load comparison of each HHV between clinical groups. The frequencies of HHV were compared using the chi□square test. For correlation analysis between HHV viral load and numerical clinical symptom scores or IgG titre from serological tests, the Spearman’s correlation was utilised. Differences where P < 0.05 are deemed significant, and in graphs * indicates P < 0.05, **P < 0.01, ***P < 0.001.

## 3 Results

### 3.1 Development of sensitive and specific ddPCR assays for human herpes viruses

For each of the nine members of the human herpes family of viruses, ddPCR assays were designed to run in duplex using specific primers and probes which were coupled to either FAM or HEX reporter labels (Supplementary Table 1). Examples of single and duplex ddPCR assays for HHV-6A and HHV-6B are shown in Figure 1A, with different proportions of positive and negative droplets reflecting the starting concentration of commercial DNA. In duplex assays spiked with both HHV-6A and HHV-6B DNA, the majority of ∼15,000 droplets measured were PCR-negative for both viruses, with substantial positivity observed for each individual virus but very few droplets deemed to have dual positivity (Figure 1B), as expected. Reproducible precise standard curves were obtained for each of the human herpes viruses (Figure 1C). Of note, the viral DNA copy number detected by ddPCR for HHV-7 and VZV was lower than the commercial DNA quantity description,: over-estimates of DNA concentration by absorbance spectrophotometry are common [29].

**Figure 1:**
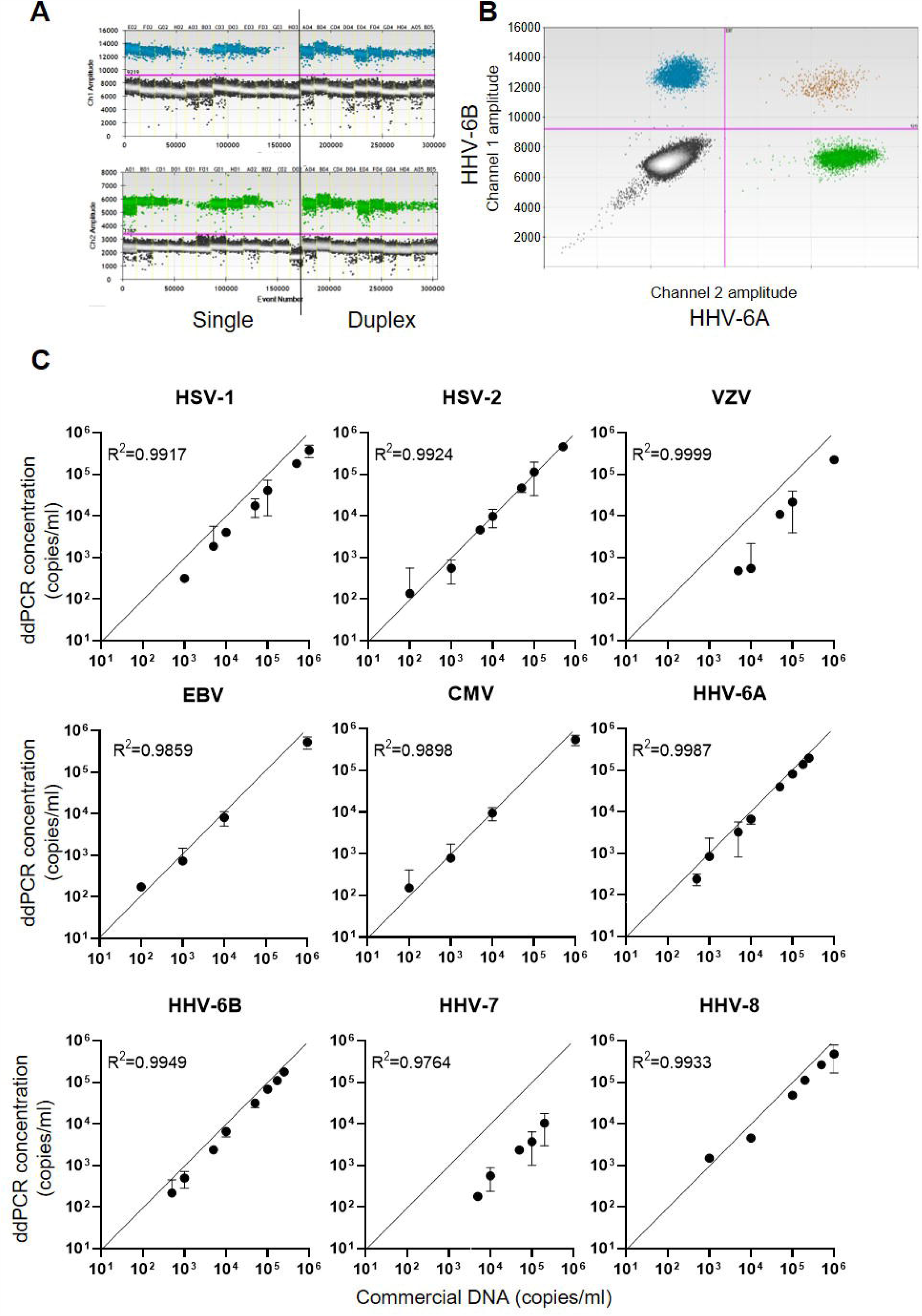
DNA concentration measured by ddPCR assays for the nine human herpes viruses. A) Example 1D dot plot data file for HHV-6A and HHV-6B ddPCR assays. The X-axes show quantities of commercial viral DNA in different assay plate wells (E02, F02, G02 etc.). Assays were run either singly or in duplex as shown. The Y-axes indicate the fluorescence intensity within each PCR droplet, with the pink line indicating the threshold for positivity. B) Example representation of 2D dot plot displaying duplex ddPCR of HHV-6B (FAM labelled, channel 1) and HHV-6A (HEX labelled, channel 2). C) Standard curves obtained for each of the nine human herpes viruses, indicating the concentration of serial dilutions of commercial DNA on the X-axes, and the calculated concentration of viral DNA from the ddPCR assay on the Y-axes.

### 3.2 Study participant selection: detection of human herpes virus DNA in plasma and PBMC

The aim of the longitudinal clinical study was to test whether the concentration of HHV DNA in saliva reflected disease severity: in order to achieve this, we needed to identify potential participants who had previously been exposed to HHVs (Figure 2). Previously [16], concentrations of IgG antibodies against HSV-1, HSV-2, VZV, EBV, CMV and HHV-6 in plasma samples from 132 people with ME/CFS, 76 HC participants and 27 people with multiple sclerosis (MS) in the UKMEB were measured by ELISA. Serum samples (189) with high IgG titre to CMV (≥ 90 U/ml) or EBV (≥ 150 U/ml for VCA or ≥90 U/ml for EBNA), or that were seropositive to ≥ five out of six HHVs measured, were screened by ddPCR to detect potential active (lytic) replication of any of the nine HHVs. HHV DNA was rarely detected, with HSV-2, HHV-6A and HHV-6B detected in plasma from one or two subjects and the remaining HHVs always negative (Table 1). We therefore screened PBMC samples, which were available from 76 potential participants; EBV was detected in 8 subjects, CMV in 1 subject and HHV-7 in 25 subjects (Table 1), probably reflecting latent infection. A single HHV DNA was detected in most cases of DNA positivity, whereas co-detection of HHVs occurred in three individuals (1 HC: CMV &HHV-7; 2 ME/CFS_SA: EBV & HHV-7) (not shown). Correlations between plasma IgG concentration and viral DNA concentration were weak due to low DNA positivity rates (data not shown). The participant characteristics are shown in Supplementary Table 2.

**Table 1.**
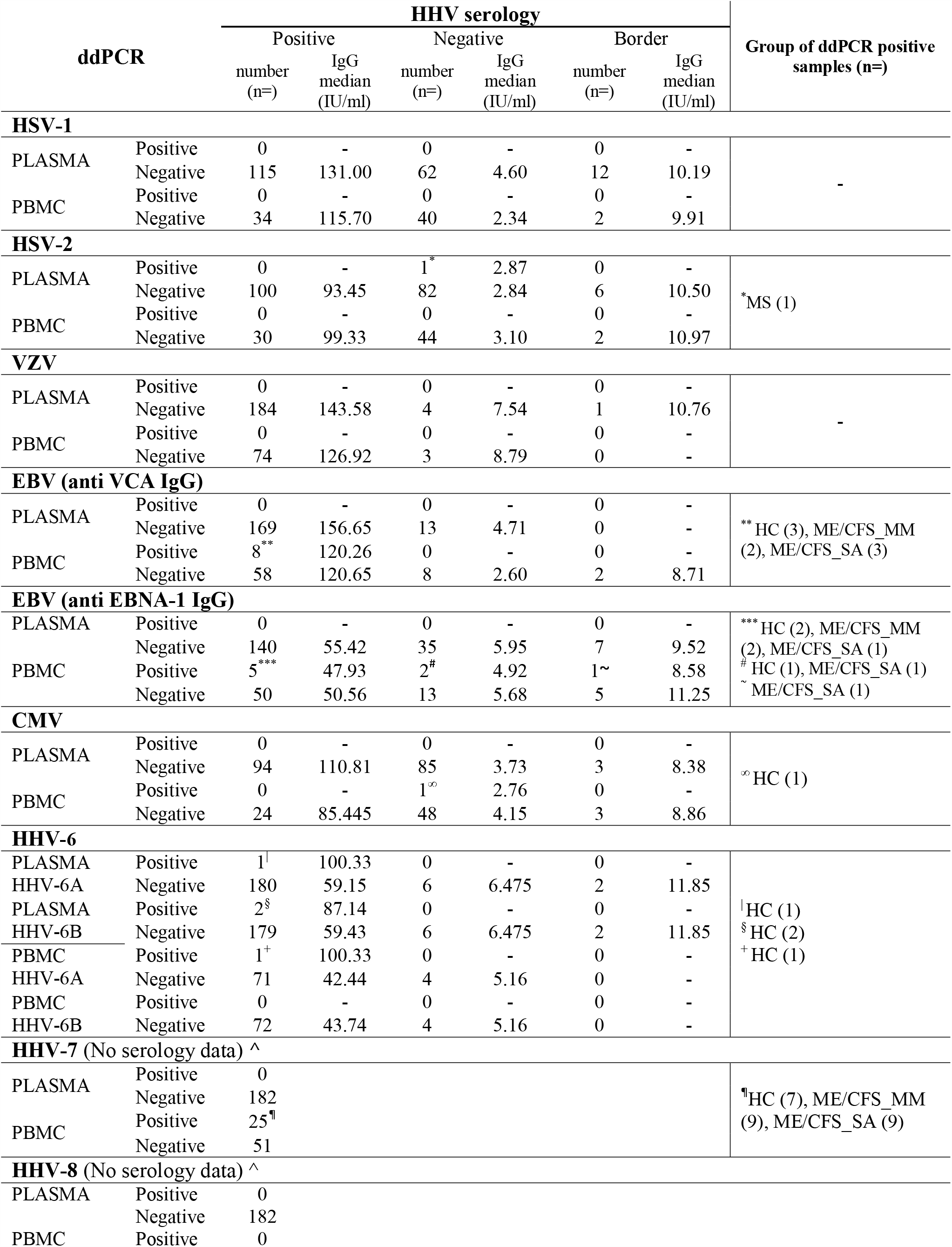
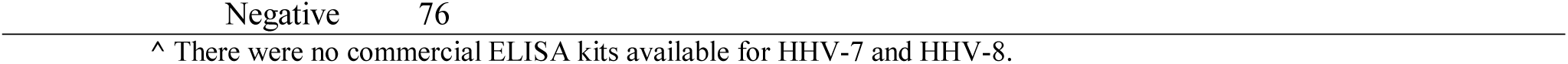
Human herpes virus DNA in plasma and PBMC with anti-HHV IgG concentration.

**Figure 2:**
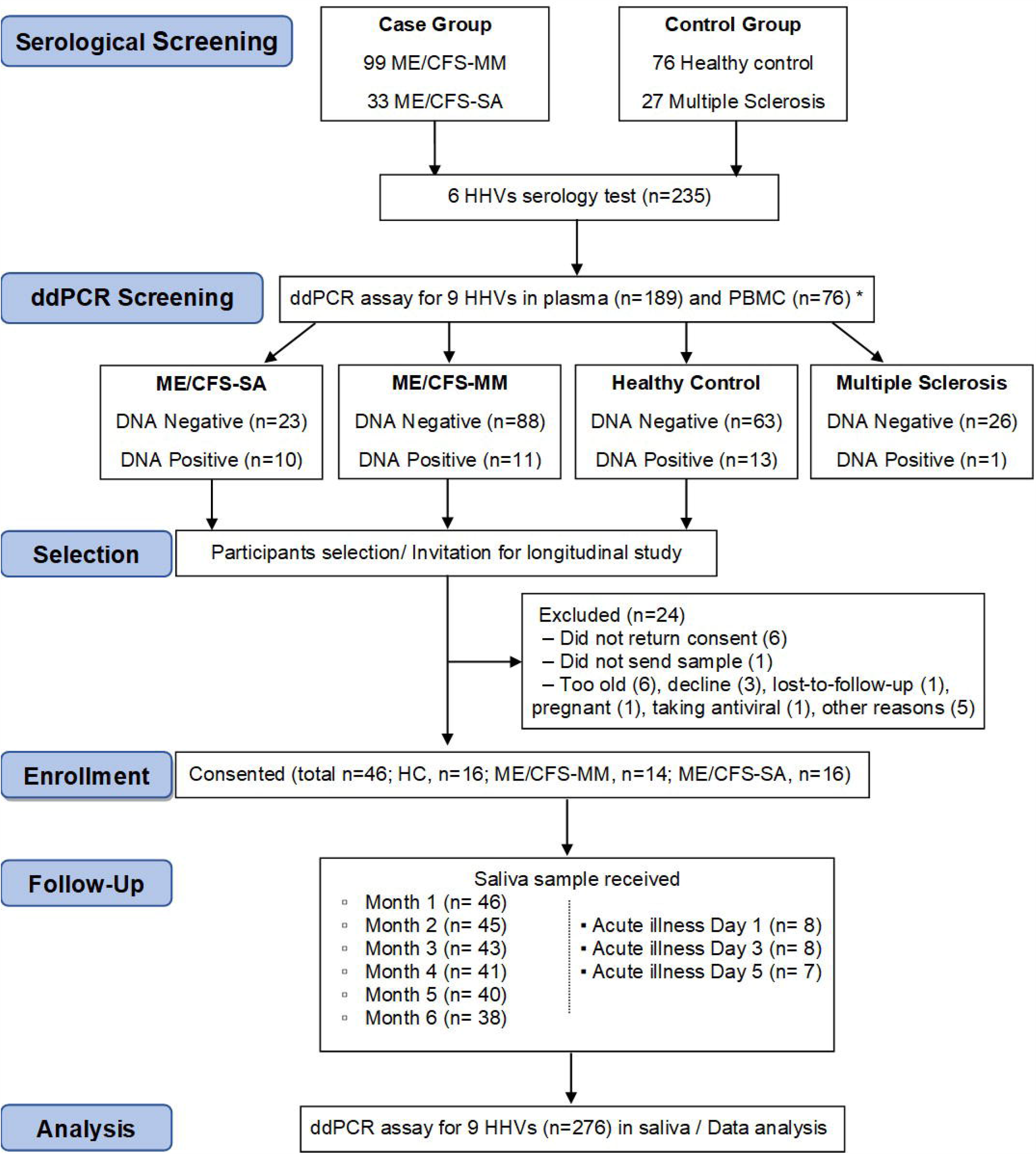
Flow diagram of participant recruitment and follow up in the longitudinal saliva clinical study. Serological test (n=235) and consecutive ddPCR assay in the plasma (n=189) and PBMC (n=76) were performed for HHV screening purposes. In total, 46 participants were enrolled into a 6-month longitudinal study to determine salivary HHV. Saliva samples (monthly and day 1,3 and 5 of an acute illness or worsening of disease episode in people with ME/CFS, n= 276) were then analysed for HHV DNA concentration by ddPCR assay, with results compared with symptoms scores. *30 participants were tested for both plasma and PBMC.

### 3.3 Longitudinal study: clinical analysis of HHVs in saliva from people living with ME/CFS

Candidates were selected for invitation to participate in the longitudinal study of saliva HHV (Figure 2) if any HHV DNA was detected in either plasma or PBMC (DNA positive) or if HHV DNA was not detected in either sample (DNA negative); all invited participants were seropositive for all six HHVs tested. MS patients, who also have samples stored in the UK ME/CFS Biobank, were not included in the longitudinal study due to low HHV DNA positivity in plasma and PBMC. A total of 70 candidates were invited to participate in the study, of whom 46 were enrolled; their blood DNA results reflected those of the group as a whole (Supplementary Table 2). Participants were requested to post a saliva sample and a completed patient questionnaire (Supplementary Figure 1) to LSHTM monthly for 6 months, and on the first, third and fifth day of any disease exacerbation episode including acute illness or significant worsening of ME/CFS symptoms. From December 2017 to June 2018, 276 saliva samples were collected. The monthly sampling return rate was 92.7 %, 83.3 %, 97.9 % in HC, ME/CFS_MM (mild/moderate) and ME/CFS_SA (severely affected) groups respectively. Additionally, one ME/CFS_MM and six ME/CFS_SA participants sent samples during episodes of acute illness/worsening of symptoms: one ME/CFS_SA sent samples for two episodes (Figure 2).

### 3.4 HHV prevalence and persistence over time in saliva from people with ME/CFS

The concentration of DNA for each of the nine HHVs was measured in saliva samples from ME/CFS patients (14 MM cases; 16 SA cases) and 16 HCs by ddPCR and the prevalence and shedding pattern of HHVs investigated over time. Throughout the study time course, HSV-1, EBV, HHV-6B and HHV-7 were detected in at least some participants, whereas the remaining HHVs were undetectable throughout. Most saliva samples contained one, two or three HHVs. At the start of the study (month 1), HHV-7 was the most prevalent HHV in saliva of all participants, whereas HSV-1 was detected from only 1 participant in each group (Figure 3A) and the prevalence of EBV and HHV-6B differed between the groups (EBV: 56% (9/16) in HC, 29% (4/14) in ME/CFS_MM, 38% (6/16) in ME/CFS_SA; HHV-6B: 25% (4/16) in HC, 29% (4/14) in ME/CFS_MM, 56% (9/16) in ME/CFS_SA). HHV prevalence remained largely unchanged throughout the 6 months of follow up with HHV-6B being the only virus to differ in prevalence between the groups, being borderline significantly more highly prevalent in severely affected ME/CFS patients than in healthy controls (Supplementary Table 3).

**Figure 3.**
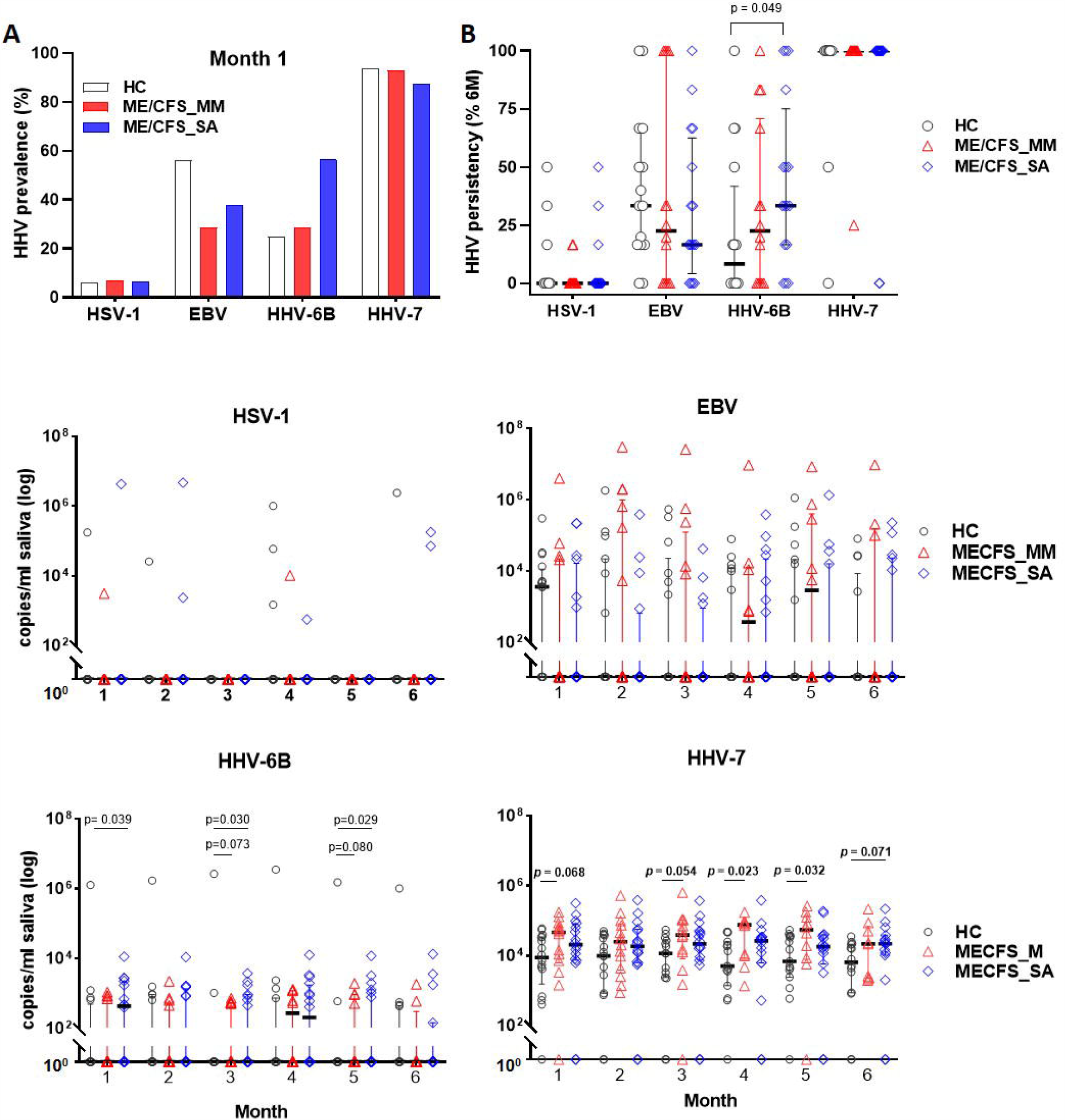
Herpes virus DNA prevalence and concentration in saliva from people living with ME/CFS, measured by ddPCR. A) Percentage HHV prevalence in HC (n=16) and people with mild/moderate ME/CFS (ME/CFS_MM: n=14) or severe symptoms (ME/CFS_SA: n=16) at month1. B) Persistence of viral DNA positivity throughout 6 months. The persistency (%) was calculated for each participant as the number of times HHV was detected divided by the total number of samples collected over 6 months, multiplied by 100. HHV viral DNA concentration comparison between HC and ME/CFS patients. Each monthly viral DNA concentration in saliva from participants with mild/moderate (ME/CFS_MM) or severe (ME/CFS_SA) disease was compared to HC across the 6 months for C) HSV-1, D) EBV, E) HHV-6B and F) HHV-7. Scatter dot plots show the median and the interquartile range. P values < 0.1 (Mann-Whitney) are shown.

To investigate viral persistence, the proportion of samples from each participant that were HHV-positive over the time course was determined (Figure 3B). HHV-7 was consistently detected in all samples from the majority of participants throughout the 6 months (median for all 3 groups = 100%), whereas HSV-1, EBV and HHV-6B were only intermittently detected. HHV-6B was detected significantly more frequently in the ME/CFS_SA participants (median: 33.3 %, IQR: 16.7-75 %), than in HC (median: 8.33 %, IQR: 0-41.7 %; p = 0.049), but not in ME/CFS_MM participants (median: 22.5 %, IQR: 0-70.8 %; p = 0.28). In contrast, there was no significant difference in the frequency of detection of EBV or HSV-1 in HC compared to people with ME/CFS (Figure 3B).

### 3.5 HHV DNA viral concentration in saliva in people living with ME/CFS

The absolute HHV viral DNA concentration load in participants with ME/CFS was compared with that in HC for the four detectable HHVs across the 6-month sampling time frame (Figure 3C-F). There were no significant differences in absolute viral DNA load for HSV (Figure 3C) or EBV (Figure 3D) between clinical groups. HHV-6B viral DNA concentrations were on average 6.2-fold higher in ME/CFS_SA patients than in HC: the differences were significant at months 1, 3 and 5, with a trend towards higher viral load across all time points (Figure 3E). There was a trend towards higher HHV-6B DNA concentration in ME/CFS-MM participants than HC, with borderline significance at months 3 and 5. Finally, HHV-7 viral loads were significantly higher in ME/CFS_MM (at months 1,3,4 and 5) and ME/CFS_SA patients (at month 6) compared to HCs, with a trend towards higher loads across all timepoints for both ME/CFS_MM and ME/CFS _SA groups, with an average 4.1-fold increase in ME/CFS-MM and 3.0-fold increase in ME/CFS_SA compared to HCs across the months (Figure 3F).

The Spearman correlation between plasma IgG concentration (in biobanked samples) and salivary HHV viral DNA load for each participant was analysed for EBV and HHV-6 (Supplementary Figure 2). There was no significant correlation between EBV viral load and either EBNA or VCA antibody concentration in people with ME/CFS or HC. Although there was a significant negative correlation between anti HHV-6 IgG concentration and salivary HHV-6B DNA concentration in the HC group, this was largely driven by one individual and no correlation was seen in the ME/CFS participants.

There was no difference in salivary HHV viral load between participants who had been deemed as HHV DNA-positive or DNA-negative in the initial plasma/PBMC ddPCR screening assay (Table 2) although, among the HC_s_, those who were plasma/PBMC HHV DNA-positive were significantly younger than those who were plasma/PBMC HHV DNA negative (Table 2). One HC in whom HHV-6B was detected in plasma had very high concentrations of HHV-6B DNA in saliva but this was an outlier and may reflect a recent infection (Supplementary Table 4). All people living with ME/CFS in whom HHV-7 was detected in PBMC also had high concentrations of salivary HHV-7 DNA (Table 2).

When stratifying participants by age, the salivary HHV-6B DNA concentration was significantly higher in younger (< 40 years) than older participants in the HC and ME/CFS_MM groups but not the ME/CFS_SA group (Supplementary Table 5) but there were no significant differences in HSV-1, EBV or HHV-7 DNA concentrations between age groups.

**Table 2:** HHV viral load in saliva of participants who were HHV DNA-positive or DNA-negative in the initial ddPCR screening assay of plasma and PBMC.

| MONTH 1 |  | N | Median Age (IQR) | p-value | Gender (n) | Median viral load (IQR, copies/ml saliva) |  |  |  |
| --- | --- | --- | --- | --- | --- | --- | --- | --- | --- |
| DNA positivity in plasma / PBMC |  |  |  |  |  | HSV-1 | EBV | HHV-6B | HHV-7 |
| HC | Negative | 11 | 46<br>(37-51) | <0.05 | F (8)/M (3) | 0 | 3600<br>(0-13,374) | 0 | 16,308<br>(682-34,600) |
|  | Positive | 5 | 32<br>(20.5-43.0) |  | F (5) | 0 | 4500<br>(0-19,017) | 0<br>(0-63700) | 10500<br>(2,400-41,167) |
| ME/<br>CFS_<br>MM | Negative | 12 | 42<br>(37.0-49.8) |  | F (12) | 0 | 0<br>(0-15580) | 0<br>(0-805) | 46,500<br>(8,192-65,350) |
|  | Positive | 2 | 47.5<br>(42.0-53.0) |  | F (2) | 0 | 30075<br>(0-60,150) | 0 | 43,653<br>(1,440-85,867) |
| ME/<br>CFS_<br>SA | Negative | 11 | 49<br>(35-54) |  | F (7)/M (4) | 0 | 0<br>(0-960) | 0<br>(0-690) | 11,400<br>(0-23183) |
|  | Positive | 5 | 35<br>(25-56) |  | F (5) | 0 | 0<br>(0-24,354) | 467.4<br>(0-2,517) | 48,300<br>(18,396-75,600) |

### 3.6 Salivary HHV viral DNA concentration through time and during acute illness episodes in people with ME/CFS

Salivary DNA concentrations of HSV-1, EBV, HHV-6B and HHV-7 varied over time in individual participants (Figure 4). Concentrations of HSV-1 and EBV DNA fluctuated month by month in the majority of infected individuals with no obvious differences between clinical groups,whereas there was less fluctuation in HHV-6B and HHV-7 DNA concentrations, with more participants with ME/CFS than HC exhibiting persistently higher viral loads (Figure 4A).

**Figure 4.**
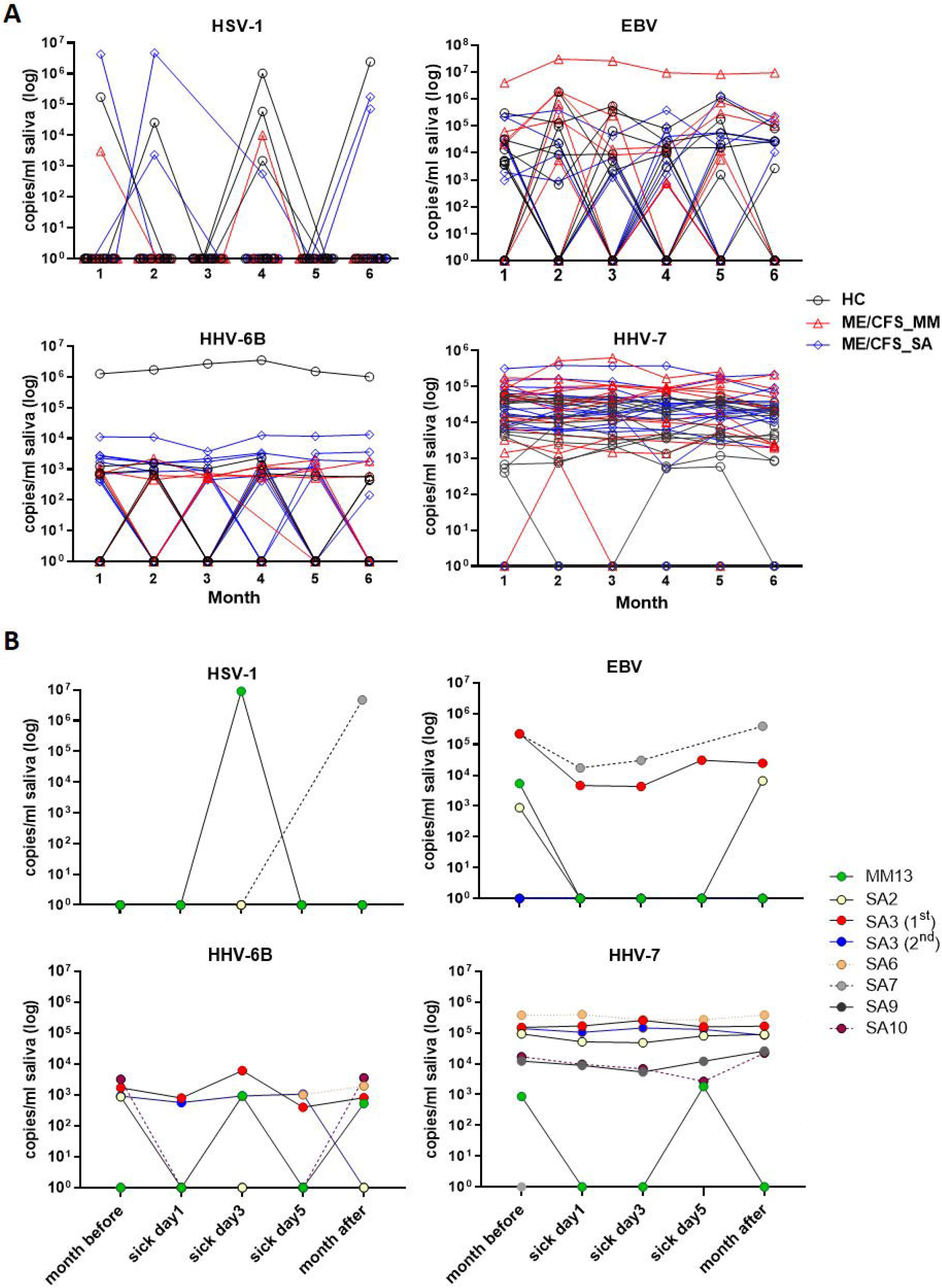
Change in herpes virus DNA concentration in saliva in individual participants over time. A) The monthly viral load of HSV-1, EBV, HHV-6B and HHV-7 detected in saliva from month 1 (Jan 2018) to month 6 (Jun 2018), measured by ddPCR assay, is shown for individual HC, ME/CFS_MM and ME/CFS_SA study participants. B) The HHV viral load of HSV-1, EBV, HHV-6B and HHV-7 in ME/CFS patients is shown on day 1, day 3 and day 5 of episodes when participants experienced an acute illness or worsening of disease symptoms.

There was no discernible pattern of changing HHV DNA concentrations during periods of worsening disease symptoms in the seven ME/CFS participants (one MM case and 6 SA, one of whom had two episodes of worsening symptoms) who had an acute illness or worsening of disease symptoms (Figure 4B): rather there was fluctuation in detection of all four viruses during these episodes, with both increases and decreases in viral load during exacerbated disease (Figure 4B and Supplementary Figure 3).

### 3.7 HHV co-infections in people with ME/CFS over time

We investigated changes in detection of viral DNA from co-infecting HHV viruses over time in individual study participants (Supplementary Figure 4 and Supplementary Table 6). At all time points, HHV-7 was more frequently detected as the only HHV present in saliva than any other HHV; HHV-7 together with HHV-6B, and HHV-7 together with EBV, as dual combinations of HHV DNAs, and the triple combination of HHV-7 together with HHV-6B and EBV, were also frequently observed (Supplementary Figure 4A). The quadruple combination (HHV-7 with HHV-6B, EBV and HSV-1) and other HHV combinations as dual or triple infections were detected only rarely (Supplementary Table 6). The HHV-7 with HHV-6B combination was significantly more frequently found in ME/CFS_SA than in HC (chi square p<0.05 at months 1 and 5), in accordance with the higher frequency of HHV-6B DNA detection in severely affected ME/CFA patients. Dual detection of HHV-7 with EBV was more frequent in ME/CFS_MM than in ME/CFS_SA participants, with significant differences (p<0.05) at months 2 and 4 and a trend in the other months.

People severely affected with ME/CFS could be broadly and evenly separated into two subgroups, dependent on how their combinations of salivary HHV DNA changed over time. People with pattern 1 showed varying combinations of one to four HHVs being detected over the 6-month time course, whereas people with pattern 2 showed relatively simpler and more stable HHV positivity consisting of one to two HHVs detectable (Supplementary Figure 4B). The majority of people with mild/moderate ME/CFS symptoms (7/10) had the more stable HHV repertoire pattern with only three ME/CFS_MM participants fitting the fluctuating repertoire pattern (data not shown).

### 3.8 Correlation of ME/CFS symptoms with each other and with viral load

To evaluate any association between disease symptoms and salivary detection of HHV DNA, the numerically graded responses to the clinical questions collected together with each saliva sample were categorised into seven domains (PEM, pain, sleep dysfunction, neurocognition, autonomic nervous system, neuroendocrine system or immune system) to yield seven “symptom scores” for each sample. First, we compared symptom scores by group (Figure 5A). HC individuals had low scores across all symptom domains. People severely affected with ME/CFS had significantly higher PEM scores (p = 0.003) and a tendency towards greater autonomic dysfunction than the ME/CFS_MM group, whereas other symptoms did not differ significantly between the two groups. We then examined the Spearman correlation of the symptom scores with each other: for all people with ME/CFS (i.e. ME/CFS_MM and ME/CFS_SA combined: n=30), all symptom scores significantly correlated with each other (median of all *r* = 0.508, p < 0.01) (Figure 5B) and symptom scores were significantly (p=0.0007) more highly correlated among patients who were mildly/moderately affected by ME/CFS than among those severely affected with ME/CFS (MM, median of *r* = 0.604; SA, median of *r* = 0.385, all p<0.05 for ME/CFS_MM group comparisons, p<0.05 for ME/CFS_SA group except Sleep dysfunction vs PEM p=0.108 and vs Pain p=0.238)(Supplementary Figure 5). Of the seven symptom domain scores, autonomic symptoms were highly correlated with neurocognition symptoms (*r* = 0.70) among all people with ME/CFS, and these two symptom domain scores were moderately correlated with other symptom scores (autonomic symptoms vs all others, median *r* = 0.586; neurocognition symptoms vs all others, median *r* = 0.582). In contrast, sleep dysfunction symptoms were only weakly correlated with other symptoms scores (median *r* = 0.348), except that among people severely affected with ME/CFS sleep dysfunction was highly correlated with neuroendocrine’ symptoms (*r* = 0.62). Finally pain symptoms were highly correlated with neurocognition, autonomic and immune related symptoms (*r* = 0.66, 0.66, 0.60, respectively) in people severely affected with ME/CFS.

**Figure 5:**
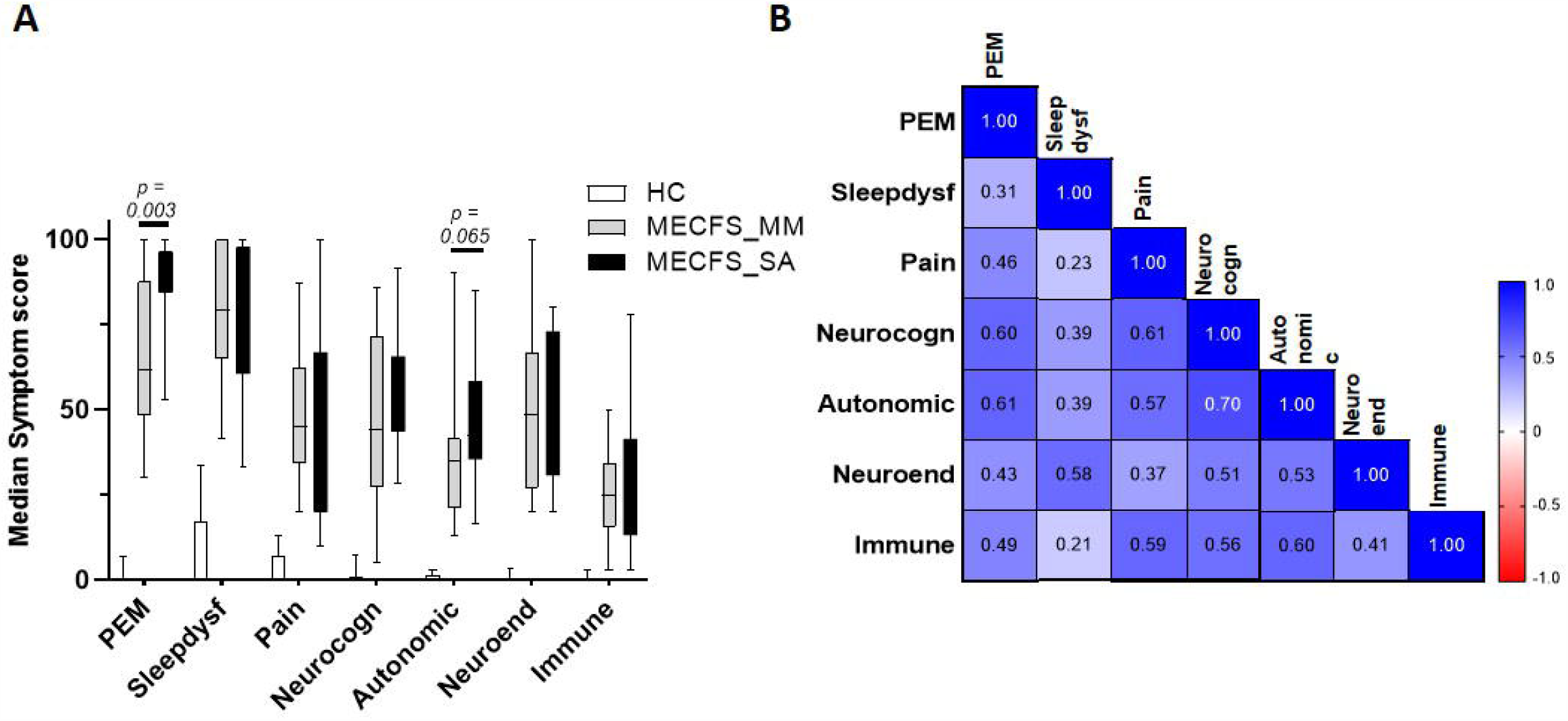
Disease symptom and correlation in people living with ME/CFS and Healthy controls. A) Median score and IQR of seven ME/CFS symptom domains across 6 months in people with mild/moderate (n=14) or severe ME/CFS (n=16) and HC (n=16), based on questionnaires completed at the time of saliva sample collection by participants. B) Correlation matrix between seven ME/CFS symptom domain scores in people with ME/CFS (n=30).

We investigated the correlation between salivary HHV DNA concentration and symptom scores across the 6 months of the study in people with ME/CFS (Supplementary Figure 5A-C) and healthy controls; overall, the extent of correlation was weak. Among people severely affected with ME/CFS, there were weak correlations between EBV DNA concentration and three symptom scores (pain, neurocognition and autonomic: r = 0.25, 0.23, 0.29 respectively, all p < 0.05), between HSV-1 DNA concentration and autonomic symptoms (r = 0.24, p < 0.05), and between HHV-7 DNA concentration and pain symptoms (r = 0.25, p <0.05) and autonomic symptoms (r = 0.24, p < 0.05). EBV DNA concentration correlated with HSV-1 DNA concentration in people severely affected with ME/CFS (r = 0.27, p < 0.01), whereas HHV-6B correlated with HHV-7 (r = 0.22, p < 0.05) (Supplementary Figure 5B). There were no significant correlations between salivary HHV DNA concentration and symptom scores among people mildly/moderately affected with ME/CFS. This group showed only weak negative correlation between EBV and HHV-6B salivary DNA concentrations (r = −0.25, p < 0.05). Among the HC there were weak but statistically significant correlations between detectable HHV-6B DNA in saliva and neuroendocrine symptoms (r = 0.21, p < 0.05) and between HSV-1 DNA concentration and both autonomic and immunological symptoms (r= 0.36, 0.22, respectively, all p < 0.05). This group also showed correlation between EBV or HHV-6B and HHV-7 salivary DNA concentrations (r = 0.28, 0.22, respectively, all p < 0.05).

We next investigated the correlation between salivary HHV DNA concentration and symptom scores in the two subgroups of severely affected ME/CFS patients (pattern 1 and pattern 2, Supplementary Figure 4B) (Figure 6, Supplementary Table 7). In the pattern 1 ME/CFS_SA group (fluctuating levels of HHV DNA in saliva) there were strong and highly significant associations between HHV-6B DNA concentration and six symptom domain scores (sleep dysfunction, pain, neurocognition, autonomic, neuroendocrine and immune, r = 0.47, 0.52, 0.65, 0.50, 0.46, 0.41, respectively, all p ≤ 0.01). Three of these symptom domains were also correlated with HHV-7 DNA concentration (pain, neurocognition and autonomic, r = 0.49, 0.44, 0.47 respectively, all p = 0.002 or< 0.001). In contrast, in the pattern 2 ME/CFS_SA group (stable HHV DNA in saliva), HHV-6B DNA concentration was negatively correlated with pain, neurocognition and neuroendocrine scores (r= −0.59, −0.43, −0.42, respectively, all p < 0.01) whereas EBV DNA concentration was positively associated with pain, neurocognition and autonomic symptoms (r = 0.48, 0.40, 0.39, respectively, p = 0.001, 0.008, 0.010 respectively). Moreover, neuroendocrine and immune system scores were significantly higher in the pattern 1 group than the pattern 2 (p < 0.01) (data not shown). Finally, when people with ME/CFS had an acute illness episode, during their sick days, the HHV-6B DNA concentrations were positively correlated with sleep dysfunction and HHV-7 DNA concentration (r=0.454, p < 0.05, r=0.548, p < 0.01 respectively; Supplementary Table 7). PEM and Pain were negatively correlated with HHV-6B, r = −0.647, p < 0.01, r = −0.426, p < 0.05 respectively).Interestingly, in all seven patients reporting an acute illness or worsening of symptoms, participants, autonomic symptoms were markedly increased whereas neuroendocrine symptoms were decreased (Supplementary Figure 6).

**Figure 6.**
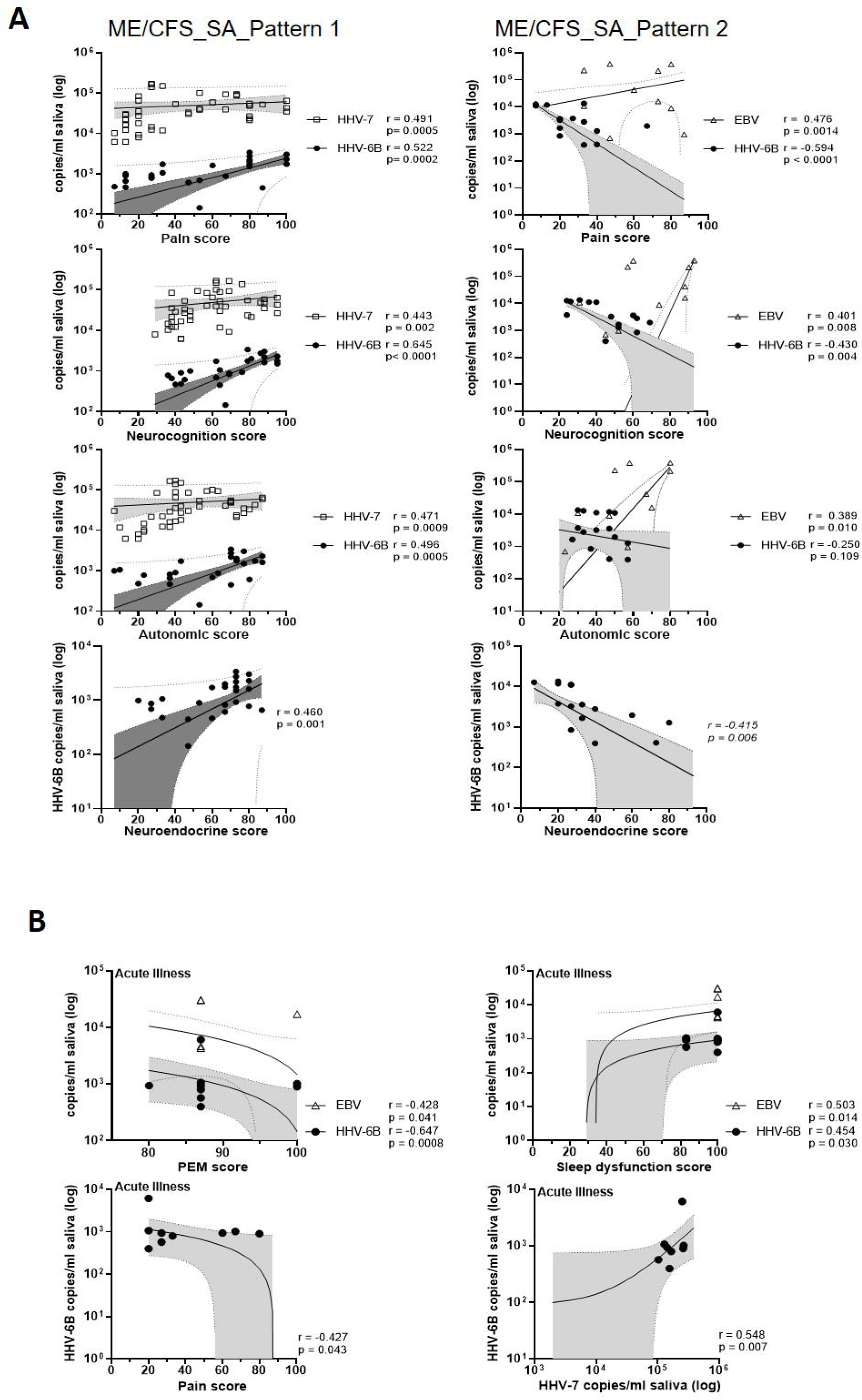
Correlations of salivary HHV DNA concentration with disease symptoms scores. A) Correlation between monthly symptoms score and monthly HHV viral DNA concentration in people with ME/CFS_SA (pattern 1, left panel; pattern 2, right panel). All participant’s monthly HHV viral DNA concentration and symptoms score data were included in the correlation analysis. Each symbol represents an individual’s monthly sample. Spearman’s coefficient is denoted as *r*. The best-fit regression line, 95% confidence interval (grey area) and prediction band (dotted line) are shown. B) Correlation between symptom scores and EBV, HHV-6B and HHV7 DNA concentrations in saliva samples from participants experiencing an exacerbation of symptoms.

## 4 Discussion

Using duplex droplet digital PCR, we have analysed the prevalence and DNA load of human herpes viruses in the saliva of ME/CFS patients and looked for associations with severity of symptoms over time. In summary, HHV-7 was the most frequently detected HHV in saliva and salivary HHV-7 DNA concentrations were significantly higher in people with ME/CFS than in healthy controls; HHV-6B was more frequently detected, and with significantly higher viral loads, in people severely affected with ME/CFS than in healthy controls and those with mild/moderate ME/CFS, such that severely affected ME/CFS patients were more likely than other groups to be HHV-6B/HHV-7 double-positive; and HHV-6B and HHV-7 viral loads correlated with disease severity in a subgroup of people severely affected with ME/CFS whose salivary HHV repertoire fluctuated over the 6 months of the study, suggesting that HHV-6B and HHV-7 may trigger ME/CFS symptoms or be reactivated in concert with worsening symptoms. Importantly, although HSV-1 and EBV were detected in some saliva samples, there were no consistent or statistically significant differences in HSV-1 and EBV reactivation between people with ME/CFS and healthy controls.

These findings are consistent with, and extend, previous data linking EBV, HHV-6 and HHV-7 infection and reactivation to the clinical course of ME/CFS (reviewed in [30]). Frequent HHV-6 and/or HHV7 infection or reactivation has been reported in chronic fatigue syndrome patients [31, 32], [33], [34] and has been linked to decreased cellular immune responses [31] and to ME/CFS symptoms including PEM and lymphadenopathy [32]. Moreover, Chu et al [35] reported that one third of patients had a documented acute HHV or B19 virus infection related to herpes viruses and B19 virus associated with ME/CFS onset.

Saliva is a key transmission route for HHVs as the salivary gland is a particularly permissive site for HHV replication. We detected HSV-1, EBV, HHV-6 and HHV-7 DNA in saliva samples, either singly or in combinations of up to 4 HHVs, consistent with previous studies [36-39]; HHV-7 has been reported to be almost universally detected in saliva [40]. The prevalence of HSV-1, EBV and HHV-7 in our healthy controls was broadly similar to a previous report Miller et al [39] but the prevalence of HHV-6B (25%) was markedly lower than the 93.5% prevalence of HHV-6 reported previously; this difference may reflect the detection target (HHV-6B rather than the whole HHV-6 genome), methodology or genuine population differences. Furthermore, in all but one of our study participants, detection of HHV DNA in saliva (evidence of virus reactivation and shedding) was highly correlated with detection of viral DNA in previously collected (baseline) samples of PBMCs (where the virus may be latent) or plasma, indicating that these are longstanding, persistent infections rather than acute/recent infections. However, in one of our healthy control participants very high plasma and salivary concentrations of HHV-6B DNA were positively correlated with immune-related and autonomic symptoms, suggestive of a primary lytic infection in this individual. Future studies could investigate correlations between viral loads in plasma, PBMCs and saliva at the same sampling time point.

Herpes viruses maintain latent infections at different anatomical sites. HHV-6 and HHV-7 remain dormant inside leukocytes, mainly T cells, and their genomes integrate into host chromosomal telomeres providing a mechanism for viral reactivation via the release of telomeric circular DNA following T cell activation in response to a heterologous infection [41, 42].Furthermore, HHV-7 can reactivate HHV-6A/B while HHV-6 has been found to activate Epstein-Barr virus from latency [43-45]. HHV-6 and HHV-7 latency within leukocytes leads to wide-ranging impacts on the immune system [4] including changes in host cell transcriptomes and metabolism [46], down-regulation of antigen presentation and modulation of cytokines and chemokines [47, 48], impairment of NK cell function [49], and enhancement of proinflammatory cytokine and chemokine responses to Toll-like receptor 9 signalling [50]. HHV-6 reactivation in ME/CFS patients can also lead to mitochondrial fragmentation and severely compromised energy metabolism [51]; this may be linked to reported interactions between the HHV-6B U95 early viral protein and the mitochondrial GRIM-19 protein [52] that results in reduced mitochondrial membrane potential and pronounced mitochondrial impairment. It has also been postulated that persistent HHV infection/reactivation during which effector T cells are controlled by regulatory T cells may also lead to ME/CFS symptoms [53]. The weak but statistically significant correlations observed here between reactivation of HHV-6, HHV-7 and EBV, and the correlations between HHV viral loads and severity of disease symptoms in ME/CFS patients, may thus reflect T cell activation either by one of these HHVs or by another infectious agent; and/or HHV-induced dysfunction of various cells of the immune system; and/or direct or indirect effects of HHVs on mitochondrial function. Whilst we did not observe any significant association between HHV reactivation and episodes of disease exacerbation, this may be due to the low number of acute disease episodes in our study, and to the marked heterogeneity of ME/CFS patients. Larger and longer studies will be needed to properly address this hypothesis and identify potential triggers.

In summary,This pilot study demonstrates that it is possible to recruit people living with ME/CFS and to collect biologically relevant samples, longitudinally, even during periods of disease exacerbation, opening up the prospect of conducting large-scale studies to investigate the role of human herpes viruses in ME/CFS pathogenesis. Our data indicate that HHV6B and HHV7 are associated with ME/CFS disease severity. Herpes virus reactivation might either be a cause or effect of ME/CFS manifestation: either a disturbance in immune system function caused by ME/CFS disease development enables a persistent HHV viral infection to become reactivated, or the reactivation of the virus might be the precipitating factor for the worsening of ME/CFS symptoms. Many factors, such as co-infection, long-term stress, immunosuppressive therapy and autoimmune disease can lead to HHV reactivation, which would fit with heterogeneity seen in ME/CFS cohorts. Long-term, large-scale studies are warranted to determine cause versus effect for the role of HHVs in ME/CFS.

## Supporting information

Combined Supplementary Data

## Data Availability

STATEMENT: The raw data supporting the conclusions of this article will be made available by the authors, without undue reservation.

## 6 Author Contributions

ER, CR, EL, LN devised the study and obtained funding, EL, CK, JN, SO conducted the clinical assessments and collected samples, JL, CR, ER, JC designed and implemented the laboratory studies JL, JC, LP, analysed data or contributed to interpretation. JL, JC, EL, CK, LP, ER, wrote the manuscript. All authors revised the manuscript critically for important intellectual content and approved the final version.

## 8 Acknowledgments

We thank all the study participants for their time and energy and for donating their regular saliva samples to this study, and their blood to the UK ME/CFS Biobank. We thank Judith Breuer for virology advice, Robert Butcher, Martin Goodier and Zakaya Nasser Al Rasbi for advice on ddPCR assay design, and Kenneth Laing for access to the droplet reader.

## 10 Data Availability Statement

The raw data supporting the conclusions of this article will be made available by the authors, without undue reservation.

