## Supplementary material for "Salivary DNA loads for human herpes viruses 6 and 7 are correlated with disease phenotype in Myalgic Encephalomyelitis/ Chronic Fatigue Syndrome": Combined Supplementary Data

### Supplementary Tables

**Supplementary Table 1. Primer and probe sequences.** Assays were performed as duplex assays by incorporation of FAM or HEX reporter labelled probes: HSV1& HSV2; EBV & CMV; HHV6A & HHV6B; HHV7 & HHV8. VZV was performed as a single assay.

| Target | Forward Primer | Reverse Primer | Probe |
|---|---|---|---|
| HSV-1 (gD_U56) | CGGCCGTGTGACACTATCG | CTCGTAAAATGGCCCCTCC | FAM-CCATACCGACCACACCGACGAACC-BHQ1 |
| HSV-2 (gG_U54) | CGCTCTCGTAAATGCTTCCCT | TCTACCCACAACAGACCCACG | HEX-CGCGGAGACATTCGAGTACCAGATCG-BHQ1 |
| VZV (DNA polymerase) | CGGCATGGCCCGTCTAT | TCGCGTGCTGCGGC | FAM-ATTCAGCAATGGAAACACACGACGCC-BHQ1 |
| EBV (BALF5) | CGGAAGCCCTCTGGACTTC | CCCTGTTTATCCGATGGAATG | FAM-TGTACACGCACGAGAAATGCGCC-BHQ1 |
| CMV (UL55) | TGGGCGAGGACAACGAA | TGAGGCTGGGAAGCTGACAT | HEX-TGGGCAACCACCGCACTGAGG-BHQ1 |
| HHV-6A (U67/68) | TTCCGGTATATGACCTTCGTAAGC | GATGTCTCACCTCCAAATCTTTAGAAAT | 6FAM-CATTATATATCGAATCTGACGCTACCTTCCG-BHQ1 |
| HHV-6B (U67/68) | | | HEX-ACATTATATGTCGAACTTGACACTACCTTCCG-BHQ1 |
| HHV-7 (gB_U39) | TTTCCTGTGACAAAAGAAGCAGTTA | ATCCCACACGCTTTACGGG | FAM-TTCCTGCGCAATAAAGTGAAAACTGTTAGCATT-BHQ1 |
| HHV-8 (MCP_Orf25) | CCACCCTCGAATGCACAAC | GTCGGGATCGGGAAAAGCT | HEX-CCACCCAGTCAGCCCAGGCACTAAAC-BHQ1 |

**Supplementary Table 2. Participants Characteristics of HHV DNA positivity from Plasma and PBMC**

| | Participant number (n=) | | Gender (n=) | Age, median (IQR) | From DNA positive sample | |
|---|---|---|---|---|---|---|
| | | | | | Detected HHV | Age range (n=) |
| **HC** | Total | 76 | F (62)/ M (14) | 44.0 (32.3-52.8) | | |
| | DNA negative | 63 | F (49)/ M (14) | 46.0 (35.0-54.0) | | |
| | DNA positive | 13 | F (13) | 32.0 (24.0-43.0) | | |
| | | | | | EBV | 18-29 (1) |
| | | | | | | 41-60 (2) |
| | | | | | HHV-6A | 30-40 (1) |
| | | | | | HHV-6B | 18-29 (1) |
| | | | | | | 30-40 (1) |
| | | | | | HHV-7 | 18-29 (3) |
| | | | | | | 30-40 (1) |
| | | | | | | 41-60 (2) |
| | | | | | CMV+HHV-7 | 30-40 (1) |
| **ME/CFS _MM** | Total | 99 | F (78)/ M (21) | 43.0 (35.0-53.0) | | |
| | DNA negative | 88 | F (68)/ M (20) | 43.5 (37.0-52.0) | | |
| | DNA positive | 11 | F (10)/ M (1) | 32.0 (25.0-53.0) | | |
| | | | | | EBV | 41-60 (2) |
| | | | | | HHV-7 | 18-29 (4) |
| | | | | | | 30-40 (2) |
| | | | | | | 41-60 (3) |
| **MC/CFS _SA** | Total | 33 | F (25) / M (8) | 48.0 (34.0-53.5) | | |
| | DNA negative | 23 | F (15) / M (8) | 49.0 (39.0-53.0) | | |
| | DNA positive | 10 | F (10) | 36.5 (30.5-55.3) | | |
| | | | | | EBV | 41-60 (1) |
| | | | | | HHV-7 | 18-29 (1) |
| | | | | | | 30-40 (4) |
| | | | | | | 41-60 (2) |
| | | | | | EBV+HHV-7 | 41-60 (1) |
| **MS** | Total | 27 | F (23) / M (4) | 52.0 (47.0-56.0) | | |
| | DNA negative | 26 | F (22) / M (4) | 51.5 (46.8-56.0) | | |
| | DNA positive | 1 | F | 53.0 (53.0-53.0) | | |
| | | | | | HSV-2 | 41-60 (1) |

**Supplementary Table 3: Prevalence of HHV positivity by time in people with ME/CFS and healthy controls.** Borderline significant results (0.05<p<0.1) are shown in **bold.**

| | | Prevalence (%) | | | Chi-square | |
|---|---|---|---|---|---|---|
| | | HC | ME/CFS_ MM | ME/CFS_ SA | *P value* HC vs MM | HC vs SA |
| **HSV-1** | M1 | 6.25 | 7.14 | 6.25 | 0.9221 | >0.999 |
| | M2 | 6.67 | 0.00 | 12.50 | 0.3255 | 0.5830 |
| | M3 | 0.00 | 0.00 | 0.00 | n/a | n/a |
| | M4 | 20.00 | 10.00 | 6.25 | 0.5040 | 0.2538 |
| | M5 | 0.00 | 0.00 | 0.00 | n/a | n/a |
| | M6 | 7.14 | 0.00 | 13.33 | 0.4123 | 0.5844 |
| **EBV** | M1 | 56.25 | 28.57 | 37.50 | 0.1269 | 0.2879 |
| | M2 | 40.00 | 42.86 | 25.00 | 0.8759 | 0.3719 |
| | M3 | 42.86 | 38.46 | 25.00 | 0.8163 | 0.3006 |
| | M4 | 40.00 | 50.00 | 43.75 | 0.6217 | 0.8325 |
| | M5 | 40.00 | 50.00 | 26.67 | 0.6217 | 0.4386 |
| | M6 | 28.57 | 33.33 | 33.33 | 0.8086 | 0.7818 |
| **HHV-6B** | M1 | 25.00 | 28.57 | 56.25 | 0.8253 | **0.0719** |
| | M2 | 33.33 | 28.57 | 37.50 | 0.7818 | 0.8085 |
| | M3 | 14.29 | 46.15 | 43.75 | **0.0700** | **0.0789** |
| | M4 | 26.67 | 50.00 | 50.00 | 0.2338 | 0.1826 |
| | M5 | 13.33 | 40.00 | 40.00 | 0.1262 | **0.0986** |
| | M6 | 28.57 | 22.22 | 26.67 | 0.7350 | 0.9087 |
| **HHV-7** | M1 | 93.75 | 92.86 | 87.50 | 0.9221 | 0.5442 |
| | M2 | 88.67 | 100.00 | 87.50 | 0.1568 | 0.9449 |
| | M3 | 85.71 | 92.31 | 87.50 | 0.5860 | 0.8859 |
| | M4 | 93.33 | 100.00 | 87.50 | 0.4047 | 0.5830 |
| | M5 | 93.33 | 90.00 | 86.67 | 0.7634 | 0.5428 |
| | M6 | 85.71 | 100.00 | 86.67 | 0.2354 | 0.9408 |

**Supplementary Table 4. HHV viral load in saliva of participants who were HHV DNA positive in the ddPCR screening assay of plasma and PBMC**

| | | | Median viral load (IQR, copies/ml saliva) for 6 months | | | |
|---|---|---|---|---|---|---|
| Participant ID (age range/gender) | | Detected HHV (sample type) | HSV-1 | EBV | HHV-6B | HHV-7 |
| **HC** | HC3 (18-29/F) | HHV-6B (plasma) | 0 | 4080 (0-60300) | 161700 (1208500-2889315) | 29348 (18095-39050) |
| | HC6 (41-60/F) | EBV (PBMC) | 0 | 0 (0-58560) | 0 | 10500 (8313-12556) |
| | HC10 (41-60/F) | HHV-6B (plasma) | 0 | 0 (0-8208) | 0 (0-108) | 0 |
| | HC12 (18-29/F) * | EBV (PBMC) | 0 | 4500 | 0 | 26333 |
| | HC13 (30-40/F) | HHV-7/CMV (PBMC) | 0 | 0 | 580 (0-790.4) | 3900 (2691-4855) |
| **ME/CFS_MM** | MM5 (41-60/F) | HHV-7 (PBMC) | 0 | 0 (0-2850) | 0 (0-660) | 261300 (129183-576350) |
| | MM10 (41-60/F) | EBV (PBMC) | 0 | 80242 (10291-197375) | 0 | 1917 (1423-3311) |
| **ME/CFS_SA** | SA11 (30-40/F) | HHV-7 (PBMC) | 0 (0-1068750) | 0 (0-1319) | 2097 (1698-2887) | 37470 (23010-50297) |
| | SA12 (30-40/F) | HHV-7 (PBMC) | 0 | 14329 (0-424833) | 0 (0-575.5) | 22500 (15583-30427) |
| | SA13 (41-60/F) | HHV-7 (PBMC) | 0 (0-19755) | 14738 (0-44250) | 72 (0-489) | 46287 (37275-61250) |
| | SA14 (41-60/F) | HHV-7/EBV (PBMC) | 0 | 0 (0-445) | 0 (0-690) | 13400 (9650-17550) |
| | SA16 (18-29/F) | HHV-7/EBV (PBMC) | 0 | 10800 (0-26152) | 2023 (1652-2847) | 50630 (36745-62950) |

*One time visited

**Supplementary Table 5. Age related HHV viral load in saliva**

| | | | Median viral load (IQR, copies/ml saliva) for 6 months | | | | | |
|---|---|---|---|---|---|---|---|---|
| | | n | EBV | P value | HHV-6B | P value | HHV-7 | P value |
| **HC** | UNDER 40 | 6 | 2040<br>(0.0-26225) | 0.696 | 290.0<br>(0-405100) | 0.036 | 27841<br>(4100 -37763) | 0.147 |
| | OVER 40 | 10 | 0.0<br>(0.0-7077) | | 0.0<br>(0.0 - 0.0) | | 7330<br>(661.3-19475) | |
| **ME/CFS_ MM** | UNDER 40 | 4 | 0.0<br>(0.0 - 0.0) | 0.296 | 591.6<br>(145.8-770.5) | 0.031 | 11571<br>(2633-35777) | 0.142 |
| | OVER 40 | 10 | 0.0<br>(0.0 - 146063) | | 0.0<br>(0.0 -0.0) | | 70375<br>(4192-103187) | |
| **ME/CFS_ SA** | UNDER 40 | 7 | 0.0<br>(0.0 - 10800) | 0.677 | 2023.0<br>(0.0-2225) | 0.132 | 23276<br>(13480 -50630) | 0.516 |
| | OVER 40 | 9 | 0.0<br>(0.0 - 7369) | | 0.0<br>(0.0 – 156.0) | | 13400<br>(2934-95087) | |

**Supplementary Table 6. Concurrent salivary HHV distribution over time**

| | HC (n=) | Month1 | Month2 | Month3 | Month4 | Month5 | Month6 |
|---|---|---|---|---|---|---|---|
| 1 | not detected | - | 1 | 1 | 1 | 1 | 1 |
| 2 | EBV only | 1 | 1 | 1 | - | - | - |
| 3 | HHV-6B ONLY | - | - | - | - | - | 1 |
| 4 | HHV-7 only | 6 | 5 | 6 | 7 | 7 | 4 |
| 5 | HHV-7+HHV-6B | - | 2 | 1 | 1 | 1 | 3 |
| 6 | HHV-7+EBV | 5 | 2 | 4 | 1 | 5 | 4 |
| 7 | HHV-7+HHV-6B+EBV | 3 | 3 | 1 | 2 | 1 | - |
| 8 | HHV-7+HHV-6B+HSV-1 | 1 | - | - | - | - | - |
| 9 | HHV-7+EBV+HSV-1 | - | - | - | 2 | - | - |
| 10 | HHV-7+HSV-1 | - | 1 | - | - | - | 1 |
| 11 | HSV-1+ EBV | - | - | - | - | - | - |
| 12 | HSV-1+EBV+HHV-6B+HHV-7 | - | - | - | 1 | - | - |
| | | 16 | 15 | 14 | 15 | 15 | 14 |

| | ME/CFS_MM (n=) | Month1 | Month2 | Month3 | Month4 | Month5 | Month6 |
|---|---|---|---|---|---|---|---|
| 1 | not detected | 1 | - | - | - | 1 | - |
| 2 | EBV only | - | - | - | - | - | - |
| 3 | HHV-6B only | - | - | 1 | - | - | - |
| 4 | HHV-7 only | 6 | 5 | 3 | 1 | 1 | 4 |
| 5 | HHV-7+HHV-6B | 3 | 3 | 4 | 2 | 3 | 2 |
| 6 | HHV-7+EBV | 2 | 5 | 4 | 4 | 4 | 3 |
| 7 | HHV-7+HHV-6B+EBV | 1 | 1 | 1 | 2 | 1 | - |
| 8 | HHV-7+HHV-6B+HSV-1 | - | - | - | 1 | - | - |
| 9 | HHV-7+EBV+HSV-1 | 1 | - | - | - | - | - |
| 10 | HHV-7+HSV-1 | - | - | - | - | - | - |
| 11 | HSV-1+EBV | - | - | - | - | - | - |
| 12 | HSV-1+EBV+HHV-6B+HHV-7 | - | - | - | - | - | - |
| | | 14 | 14 | 13 | 10 | 10 | 9 |

| | ME/CFS_SA (n=) | Month1 | Month2 | Month3 | Month4 | Month5 | Month6 |
|---|---|---|---|---|---|---|---|
| 1 | not detected | 1 | 1 | 1 | 1 | 1 | 1 |
| 2 | EBV only | 1 | - | 1 | - | 1 | - |
| 3 | HHV-6B only | - | - | - | - | - | - |
| 4 | HHV-7 only | 4 | 6 | 6 | 5 | 4 | 7 |
| 5 | HHV-7+HHV-6B | 5 | 4 | 5 | 3 | 6 | 2 |
| 6 | HHV-7+EBV | 1 | 1 | 1 | 1 | 3 | 2 |
| 7 | HHV-7+HHV-6B+EBV | 3 | 2 | 2 | 5 | - | 1 |
| 8 | HHV-7+HHV-6B+HSV-1 | 1 | - | - | - | - | - |
| 9 | HHV-7+EBV+HSV-1 | - | - | - | - | - | - |
| 10 | HHV-7+HSV-1 | - | 1 | - | - | - | - |
| 11 | HSV-1+EBV | - | 1 | - | 1 | - | 1 |
| 12 | HSV-1+EBV+HHV-6B+HHV-7 | - | - | - | - | - | 1 |
| | | 16 | 16 | 16 | 16 | 15 | 15 |

**Supplementary Table 7. The R value of Spearman's analysis between HHV virus and ME/CFS symptom scores from ME/CFS_SA**

| Spearman r | | HSV-1 | EBV | HHV-6B | HHV-7 | PEM | Sleepdysf | Pain | Neurocogn | Autonomic | Neuroend | Immune |
|---|---|---|---|---|---|---|---|---|---|---|---|---|
| **ME/CFS_SA-pattern 1 Fluctuating repertoire (n = 8)** | HSV-1 | 1 | | | | | | | | | | |
| | EBV | -0.033 | 1 | | | | | | | | | |
| | HHV-6B | 0.055 | 0.096 | 1 | | | | | | | | |
| | HHV-7 | 0.102 | 0.253 | 0.167 | 1 | | | | | | | |
| | PEM | 0.232 | -0.022 | 0.284 | 0.202 | 1 | | | | | | |
| | Sleepdysf | -0.122 | -0.182 | $0.472^{***}$ | -0.202 | 0.120 | 1 | | | | | |
| | Pain | 0.158 | 0.100 | $0.522^{***}$ | $0.491^{***}$ | $0.646^{***}$ | -0.010 | 1 | | | | |
| | Neurocogn | 0.117 | 0.046 | $0.650^{***}$ | $0.443^{**}$ | $0.414^{**}$ | $0.308^{*}$ | $0.746^{***}$ | 1 | | | |
| | Autonomic | 0.168 | 0.205 | $0.496^{***}$ | $0.472^{***}$ | $0.524^{***}$ | 0.038 | $0.835^{***}$ | $0.666^{***}$ | 1 | | |
| | Neuroend | -0.034 | -0.180 | $0.460^{**}$ | 0.053 | 0.105 | $0.724^{***}$ | 0.147 | 0.263 | $0.294^{*}$ | 1 | |
| | Immune | 0.243 | -0.021 | $0.407^{**}$ | 0.060 | $0.480^{***}$ | 0.156 | $0.719^{***}$ | $0.410^{**}$ | $0.762^{***}$ | $0.335^{*}$ | 1 |
| **ME/CFS_SA-pattern 2 Stable repertoire (n = 7)** | HSV-1 | 1 | | | | | | | | | | |
| | EBV | $0.597^{***}$ | 1 | | | | | | | | | |
| | HHV-6B | -0.200 | -0.261 | 1 | | | | | | | | |
| | HHV-7 | $-0.412^{**}$ | $-0.453^{**}$ | 0.176 | 1 | | | | | | | |
| | PEM | -0.016 | 0.288 | -0.240 | $-0.324^{*}$ | 1 | | | | | | |
| | Sleepdysf | 0.146 | 0.256 | -0.033 | -0.103 | 0.251 | 1 | | | | | |
| | Pain | 0.160 | $0.476^{**}$ | $-0.594^{***}$ | -0.055 | $0.467^{**}$ | $0.322^{*}$ | 1 | | | | |
| | Neurocogn | 0.260 | $0.401^{**}$ | $-0.431^{**}$ | $-0.306^{*}$ | $0.358^{*}$ | 0.213 | $0.557^{***}$ | 1 | | | |
| | Autonomic | $0.316^{*}$ | $0.389^{*}$ | -0.250 | -0.170 | 0.246 | $0.697^{***}$ | $0.354^{*}$ | $0.429^{**}$ | 1 | | |
| | Neuroend | 0.005 | 0.262 | $-0.415^{**}$ | -0.146 | $0.549^{***}$ | $0.538^{***}$ | $0.624^{***}$ | $0.556^{***}$ | $0.611^{***}$ | 1 | |
| | Immune | 0.088 | 0.183 | -0.157 | -0.145 | $0.388^{*}$ | 0.169 | $0.471^{**}$ | $0.590^{***}$ | 0.298 | $0.388^{*}$ | 1 |
| **ME/CFS_ acute illness episode (ME/CFS_SA; n=6, ME/CFS_MM; n=1)** | HSV-1 | 1 | | | | | | | | | | |
| | EBV | -0.111 | 1 | | | | | | | | | |
| | HHV-6B | 0.292 | 0.139 | 1 | | | | | | | | |
| | HHV-7 | -0.306 | 0.003 | $0.548^{**}$ | 1 | | | | | | | |
| | PEM | -0.409 | $-0.428^{*}$ | $-0.647^{**}$ | -0.089 | 1 | | | | | | |
| | Sleepdysf | 0.205 | $0.504^{*}$ | $0.454^{*}$ | 0.211 | -0.321 | 1 | | | | | |
| | Pain | -0.016 | -0.203 | $-0.426^{*}$ | -0.226 | $0.625^{**}$ | 0.258 | 1 | | | | |
| | Neurocogn | 0.325 | 0.112 | 0.059 | -0.382 | -0.294 | 0.314 | 0.239 | 1 | | | |
| | Autonomic | -0.179 | 0.204 | -0.195 | 0.019 | 0.300 | 0.381 | 0.380 | 0.130 | 1 | | |
| | Neuroend | -0.260 | 0.306 | 0.268 | 0.315 | -0.066 | $0.526^{*}$ | 0.133 | 0.070 | $0.517^{*}$ | 1 | |
| | Immune | 0.228 | -0.384 | -0.122 | -0.043 | $0.597^{**}$ | 0.140 | $0.564^{**}$ | -0.063 | 0.376 | 0.129 | 1 |

**Supplementary Figures**

**ME/CFS RESEARCH PROJECT**
**SYMPTOMS ASSESSMENT – DAY 1 of acute illness or significant worsening of symptoms**

| Sex: Male ☐ Female ☐ | Project ID Nº (For LSHTM use only) |
| --- | --- |
| Date of birth: ____/____/_______ (dd/mm/yyyy) | Today's date: ____/____/_______ (dd/mm/yyyy) |

A. Please describe your current illness or worsening of symptoms.

__________________________________________________
__________________________________________________
__________________________________________________
__________________________________________________

1. How much time during the past week...

| | None of the time | A little of the time | Some of the time | A good bit of the time | Most of the time | All of the time |
| --- | --- | --- | --- | --- | --- | --- |
| Did you feel worn out? | 0 | 1 | 2 | 3 | 4 | 5 |
| Did you have a lot of energy? | 0 | 1 | 2 | 3 | 4 | 5 |
| Did you feel tired? | 0 | 1 | 2 | 3 | 4 | 5 |
| Did you have enough energy to do the things you wanted to do? | 0 | 1 | 2 | 3 | 4 | 5 |
| Did you feel full of pep (lively)? | 0 | 1 | 2 | 3 | 4 | 5 |

2. Please mark the line with an 'X' to describe the severity of pain that you are currently experiencing (think of the last week including today), from no pain at all (mark on the left of the line) to maximum pain (mark on the right of the line).

**No pain** ____________________________________ **Pain as bad as possible**

3. Please mark the line with an 'X' to describe the severity of fatigue that you are currently experiencing (think of the last week including today), from no fatigue at all (mark on the left of the line) to maximum fatigue (mark on the right of the line).

**No fatigue** ____________________________________ **Fatigue as bad as possible**

Please mark the next two lines thinking about how much of that fatigue is physical and how much is mental.

**No physical fatigue** ____________________________________ **Physical fatigue as bad as possible**

**No mental fatigue** ____________________________________ **Mental fatigue as bad as possible**

4. Please tick the boxes using the scale above them to tell us more about which of the symptoms listed below you have had over the past week:

| | Absent | Mild | Moderate | Severe |
| --- | --- | --- | --- | --- |
| 1. Sore throat | | | | |
| 2. Symptoms of flu | | | | |
| 3. Fever or chills | | | | |
| 4. Tender glands in the neck or arm pit | | | | |
| 5. Viral infections | | | | |
| 6. Sensitivities to food, medications, chemicals, smells/odours, and/or other | | | | |
| 6.a. Intolerance to alcohol (alcohol makes you feel ill/much worse) | | | | |
| 7. Feeling ill (malaise) after exertion/activity which lasts for a long time (more than 24 hours) | | | | |
| 8. Pain after exertion/activity which lasts for a long time (more than 24 hours) | | | | |
| 9. Muscle pain | | | | |
| 10. Muscle discomfort | | | | |
| 10.a. Muscle twitching | | | | |
| 11. Stiffness in the mornings | | | | |
| 11a. Pain in chest or abdomen | | | | |
| 12. Pain in two or more joints, without swelling or redness | | | | |
| 13. Joint pains moving to different joints without swelling or redness | | | | |

| |
|---|
| 14. Neck weakness |
| 15. Back weakness |
| 16. Air hunger, difficulty in breathing or shortness of breath on exertion/activity |
| 17. Muscle weakness |
| 18. Headaches which are new, different or worse than before the disease started |
| 19. Migraine which is different/worse than before |
| 20. Unusual sensitivity to light and/or noise |
| 21. Temporary disturbance in eyesight |
| 22. Tingling or numbness in arms and/or legs |
| 23. Loss of balance, unsteadiness on feet when standing, or inability to focus the vision |
| 24. Poor coordination or unsteady movements (unsteadiness on walking) |
| 25. Short term memory problems |
| 26. Trouble concentrating |
| 27. Confusion or 'brain fog' |
| 28. Disorientation |
| 29. Difficulty understanding things, thinking clearly |
| 29.a.Difficulty finding or saying words |
| 30. Difficulty retaining/ recalling information |
| 31. Slow thinking |
| 31a. Difficulty making decisions |
| 32. Unrefreshing sleep |
| 33. Problems in sleep quality or duration (other than sleep apnoea), such as insomnia, changing night for day, awakening during the night |
| 34. Marked physical or mental fatigue or exhaustion after **minimal** exertion/activity, which lasts for a long time (more than 24 hours) |
| 35. Fatigue or exhaustion after levels of activity that should normally not cause fatigue (i.e. would not cause fatigue before you got ill) |
| 36. Intolerance to exercise (it is difficult to exercise) |
| 37. Worsening of symptoms after exertion/activity, which lasts for a long time (more than 24 hours) |
| 38. Worsening of symptoms with stress |

Page 3

| |
|---|
| 39. Intolerance to (difficulty with) standing on your feet |
| 40. Dizziness while standing up |
| 41. Palpitations while **standing up** |
| 41a. Palpitations (feeling that your heart is racing or pounding) |
| 42. Feeling light-headed |
| 43. Being extremely pale |
| 44. Being unusually sweaty |
| 44a. Unusually cold hands or feet |
| 45. Intolerance to extremes of heat and cold |
| 46. Feeling sick (nausea) |
| 46. a. Irritable bowel symptoms, such as diarrhoea or constipation, abdominal pain, and bloating |
| 47. Bladder problems, such as having a sudden need to urinate (pass water), urinating more often than usual, or waking up during the night to pass water |
| 48. Decreased sexual interest and/or function |
| 49. Abnormal appetite or significant changes in weight (not intentional) |

END OF FORM - **If you have any queries, please phone the research nurse on 020 7XXX XXXX and she will be happy to help you.**

*Thank you for continuing to participate!*

Page 4

**Supplementary Figure 1. Clinical symptom assessment forms.** The form was completed by the participant at the time that each saliva sample was collected and returned, i.e. at Month 1, 2, 3, 4, 5 and 6 and then there was an disease exacerbation episode.

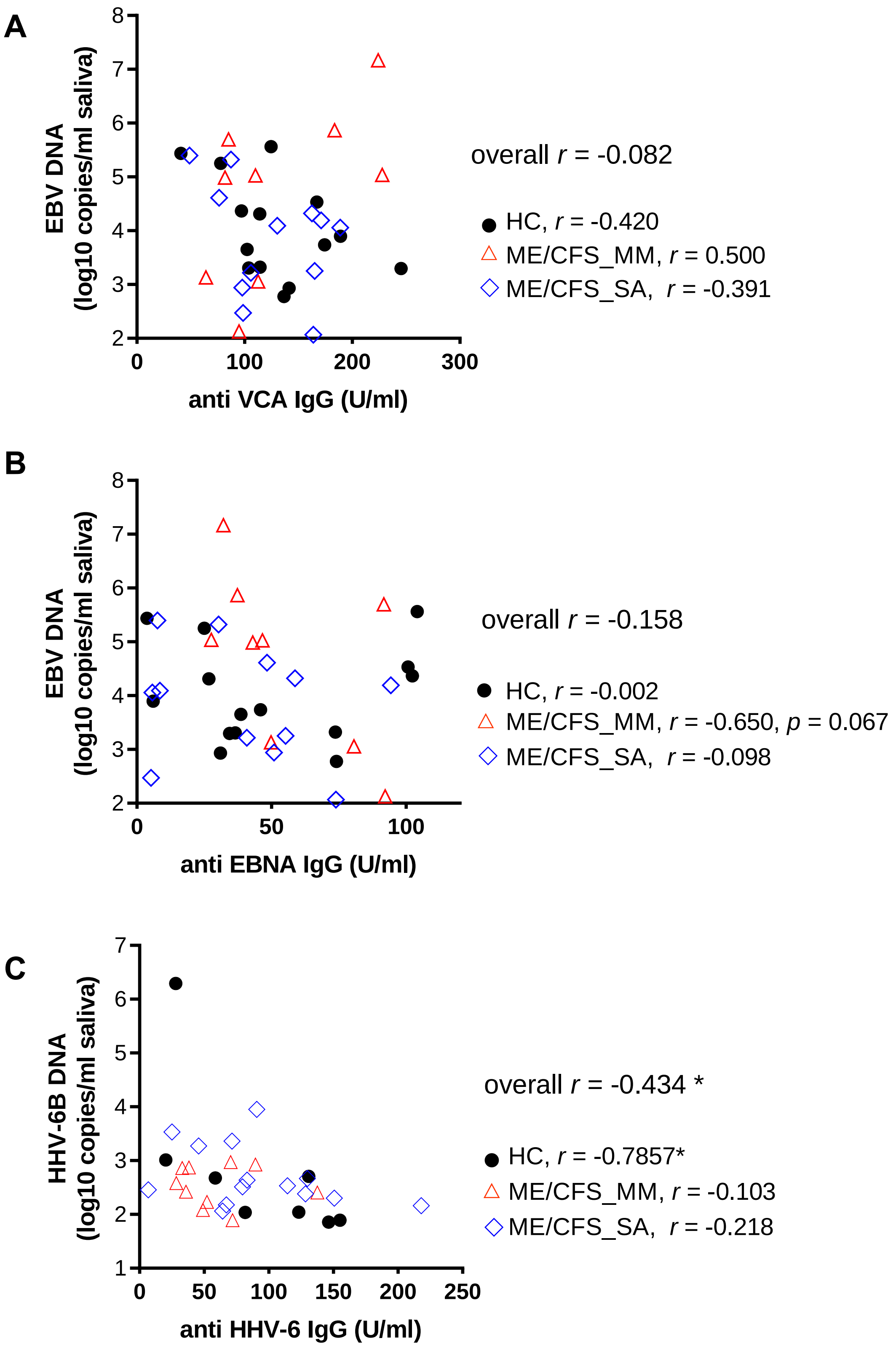

**Supplementary Figure 2. Correlation analysis between salivary HHV viral DNA concentration and anti HHV IgG titre.** (A) EBV viral DNA concentration with anti VCA IgG (B) EBV viral DNA concentration with anti EBNA IgG (C) HHV-6B viral load with anti HHV-6 IgG. Average viral DNA concentration measured monthly for 6 months was analysed with previously measured plasma anti HHV IgG level. Spearman's coefficient was denoted as *r*. *, $p < 0.05$

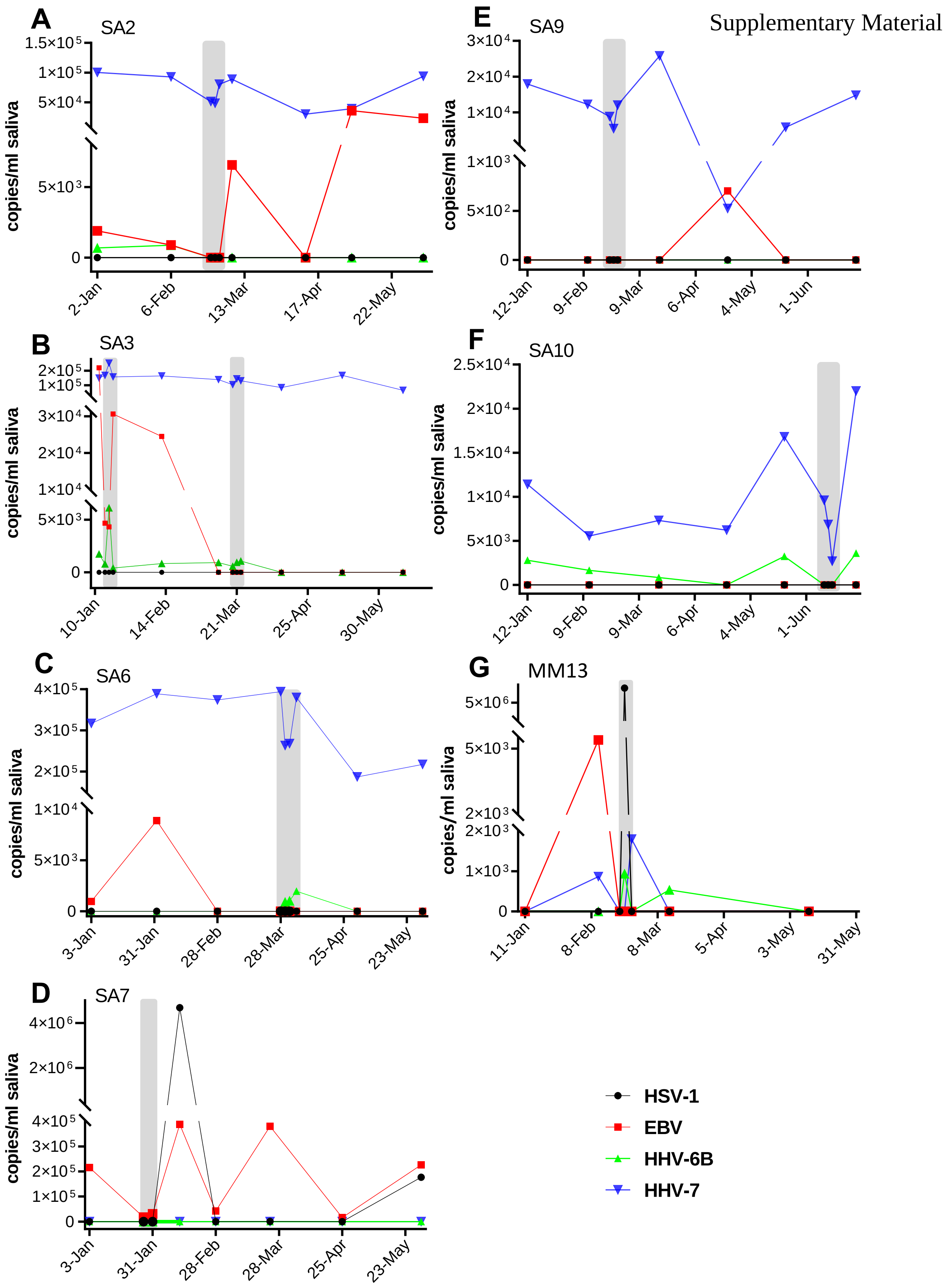

**Supplementary Figure 3. Change of HHV viral DNA concentrations in people with ME/CFS (n=7) who had acute illness or worsening disease severity** (highlighted in grey).

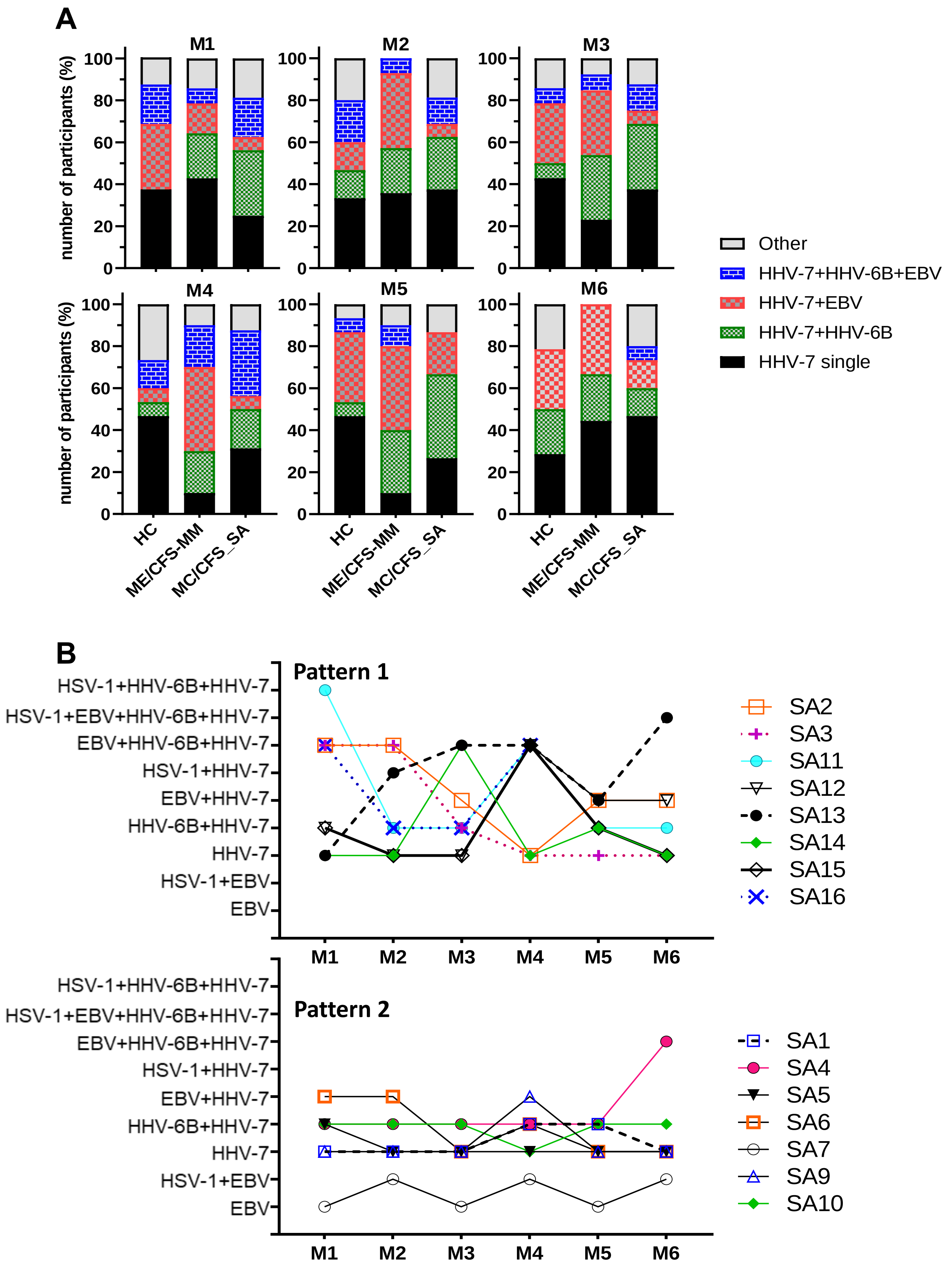

**Supplementary Figure 4. Concurrent salivary HHV distribution over time**. A) Distribution of different combinations of HHV DNA detected in saliva from participants with mild/moderate (MM) or severe (SA) ME/CFS and healthy controls (HC). B) Severely affected ME/CFS patients had either fluctuating HHV DNA repertoires (Pattern 1) or more constant repertoires (Pattern 2).

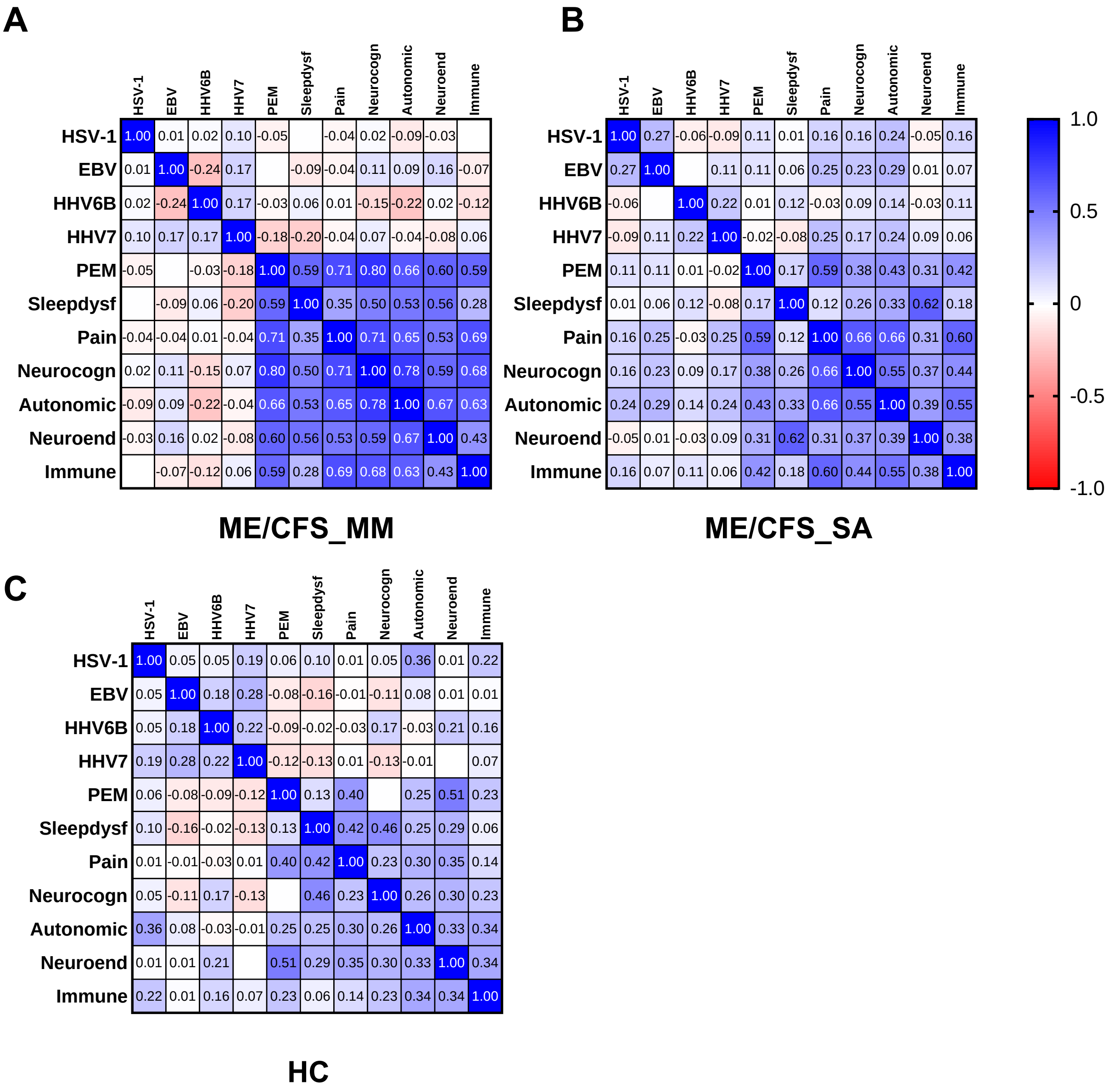

**Supplementary Figure 5. Correlation matrixes of seven typical symptoms domains of ME/CFS with HHV viral DNA concentration in saliva from people with ME/CFS.** Correlation efficient between HHV viral loads and symptoms scores across 6 months in (A) ME/CFS_MM, (B) ME/CFS_SA and HC (C)

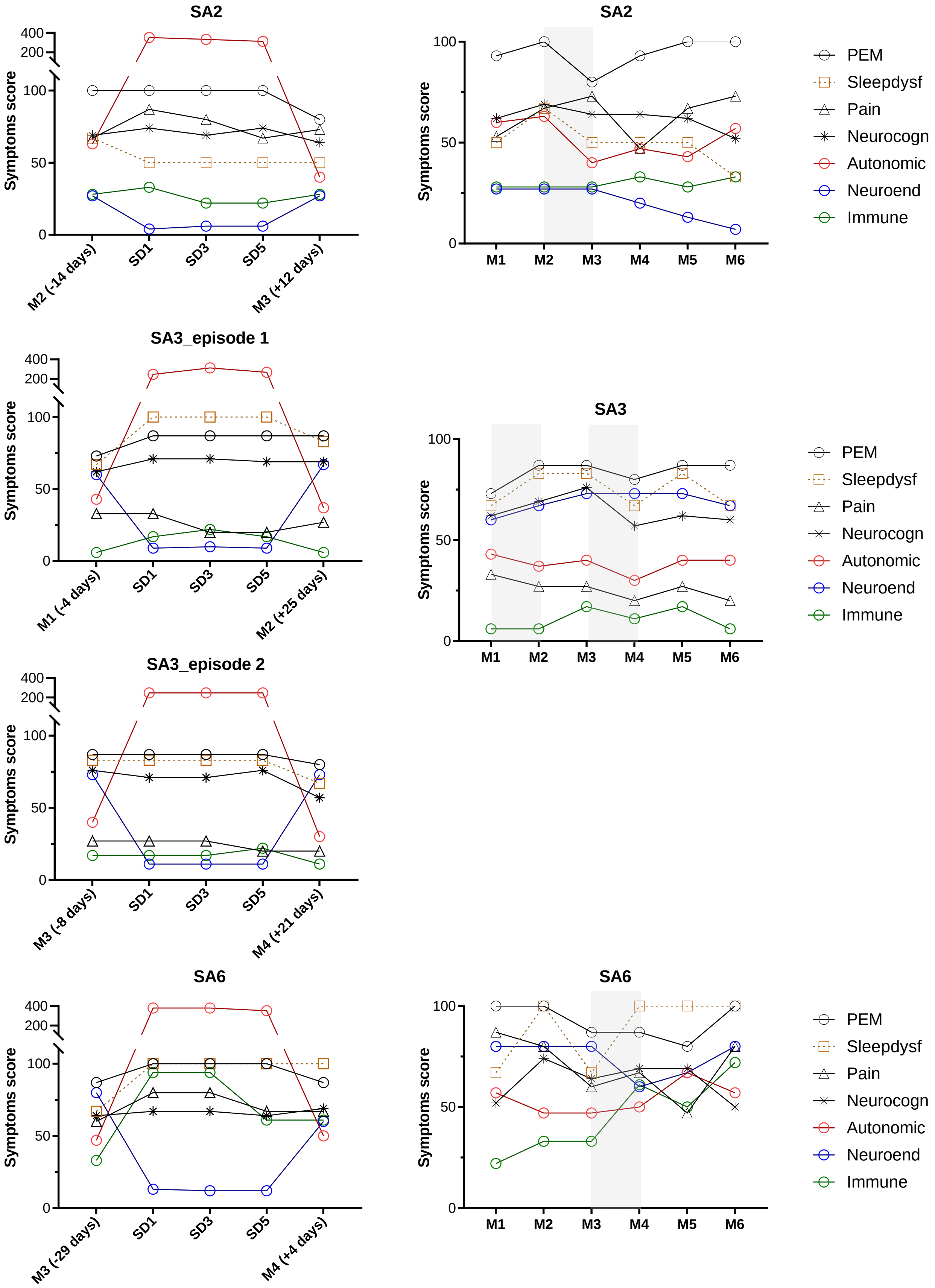
SA2
Symptoms score
M2 (-14 days)
SD1
SD3
SD5
M3 (+12 days)
SA2
Symptoms score
M1
M2
M3
M4
M5
M6
PEM
Sleepdysf
Pain
Neurocogn
Autonomic
Neuroend
Immune
SA3_episode 1
Symptoms score
M1 (-4 days)
SD1
SD3
SD5
M2 (+25 days)
SA3
Symptoms score
M1
M2
M3
M4
M5
M6
PEM
Sleepdysf
Pain
Neurocogn
Autonomic
Neuroend
Immune
SA3_episode 2
Symptoms score
M3 (-8 days)
SD1
SD3
SD5
M4 (+21 days)
SA6
Symptoms score
M3 (-29 days)
SD1
SD3
SD5
M4 (+4 days)
SA6
Symptoms score
M1
M2
M3
M4
M5
M6
PEM
Sleepdysf
Pain
Neurocogn
Autonomic
Neuroend
Immune

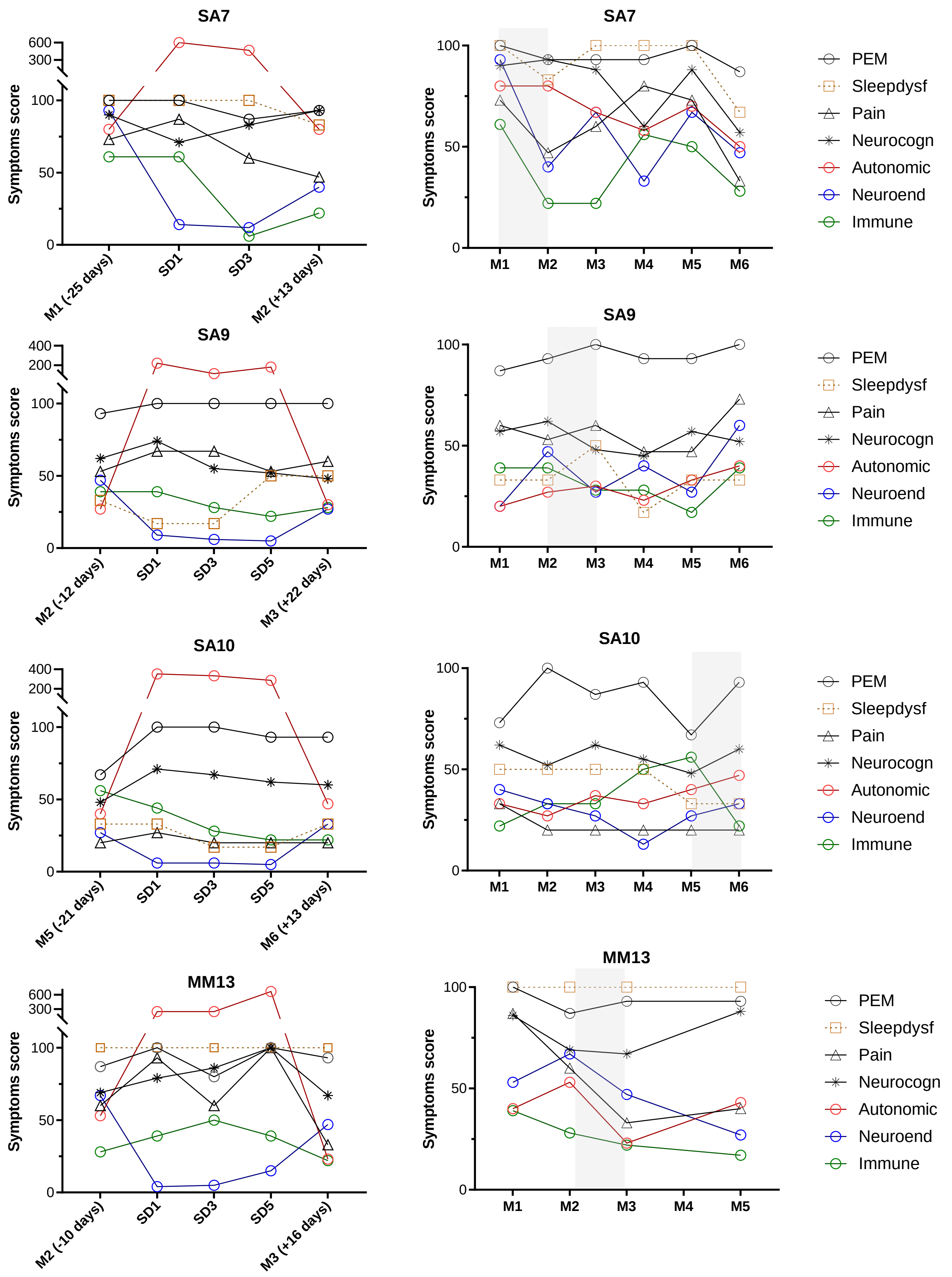

**Supplementary Figure 6. Change of symptoms score in people with ME/CFS (n=7) who had acute illness or worsening of disease symptoms**. Sick days (left panel) and monthly (right panel). Sick days occurred in the light grey area.
